# Estimating the mpox vaccine uptake among MSM and modelling the potential of future vaccination campaigns in the EU/EEA

**DOI:** 10.64898/2026.04.16.26350851

**Authors:** Bastian Prasse, Disa Hansson, Liana Aphami, Kai J. Jonas, Jordi Borrel Pique, Xanthi Andrianou, Anastasia Pharris, Diamantis Plachouras, Axel J. Schmidt, Lina Nerlander

## Abstract

In October 2025, mpox virus clade I infections have been detected among men who have sex with men (MSM) in the EU/EEA, suggesting local transmission in MSM sexual networks. Given the large outbreak of mpox among MSM in 2022 and the uncertain transmission parameters of clade I in the European context, clade I poses a public health concern to the EU/EEA. This work assesses the potential effect of increasing the mpox vaccine uptake among MSM via two contributions. First, building on the European MSM and Trans Persons Internet Survey 2024, we estimate the mpox vaccine uptake among MSM as well as the proportion who are unvaccinated but willing to get vaccinated for 28 countries in the EU/EEA. Specifically, we fit Bayesian mixed-effects models for the vaccine and recovery status of an individual depending on their number of sexual partners and country. Second, we develop a susceptible-infectious-recovered model on a sexual contact network to estimate the reduction of the reproduction number if vaccines are provided to MSM who are willing to get vaccinated. Our results suggest a substantial willingness for mpox vaccination among MSM if mpox cases increase and a large reduction of the effective reproduction number if this willingness is met. These findings highlight a large potential of increasing mpox vaccine uptake among MSM and preventing future mpox outbreaks in the EU/EEA.

## Introduction

Mpox is a zoonotic orthopoxvirus infection first identified in humans in 1970 in the Democratic Republic of the Congo (DRC). Cases remained limited to sporadic outbreaks until 2022, when substantial global transmission prompted the World Health Organization (WHO) to declare a Public Health Emergency of International Concern (PHEIC) [1]. In the European Union and the European Economic Area (EU/EEA), more than 21,000 cases were reported to ECDC between May and November 2022, most of which were among men who have sex with men (MSM) [2, 3]. Since the end of 2022, the mpox incidence in the EU/EEA has remained stable at low levels due to a combination of factors, including behavioural change, accumulation of natural immunity through recovery, vaccination campaigns, and contact tracing efforts [4-6].

There are two clades of mpox virus (MPXV), clade I and clade II. The 2022 outbreak that triggered the PHEIC was due to MPXV clade II, and the WHO declared the PHEIC to be over in 2023. In August 2024, WHO declared a second PHEIC due to the rapid increase of MPXV clade Ib cases in the DRC and neighbouring countries [7]. Between August 2024 and October 2025, there were occasional travel-associated MPXV clade Ib cases in the EU/EEA, with limited secondary transmission. In October 2025, MPXV clade Ib cases were detected in MSM sexual networks in the EU/EEA without any travel links, indicating ongoing local transmission [8].

Compared to the outbreak in 2022, the mpox notifications of clade II aggregated across the EU/EEA has been relatively constant in 2024 and 2025. However, there have been marked fluctuations of the mpox notifications on country-level, suggesting that mpox circulation in the EU/EEA is driven by consecutive regional outbreaks in different, possibly overlapping, MSM sexual networks [3]. Figure 1 suggests that a growing proportion of mpox cases in the EU/EEA seems to be due to MPXV clade I rather than MPXV clade II – however, the epidemiological situation is evolving and there are substantial uncertainties. Current observations from surveillance data do not indicate differences in severity between clade I and clade II in the EU/EEA context, however, strong evidence on clade I parameters in the EU/EEA context is missing [8]. Furthermore, the fact that two different MPXV clades caused major outbreaks highlights the need to be prepared against potential new future MPXV (sub-)clades, to which this work contributes by studying the potential of increasing the vaccine uptake among MSM.

**Figure 1.**
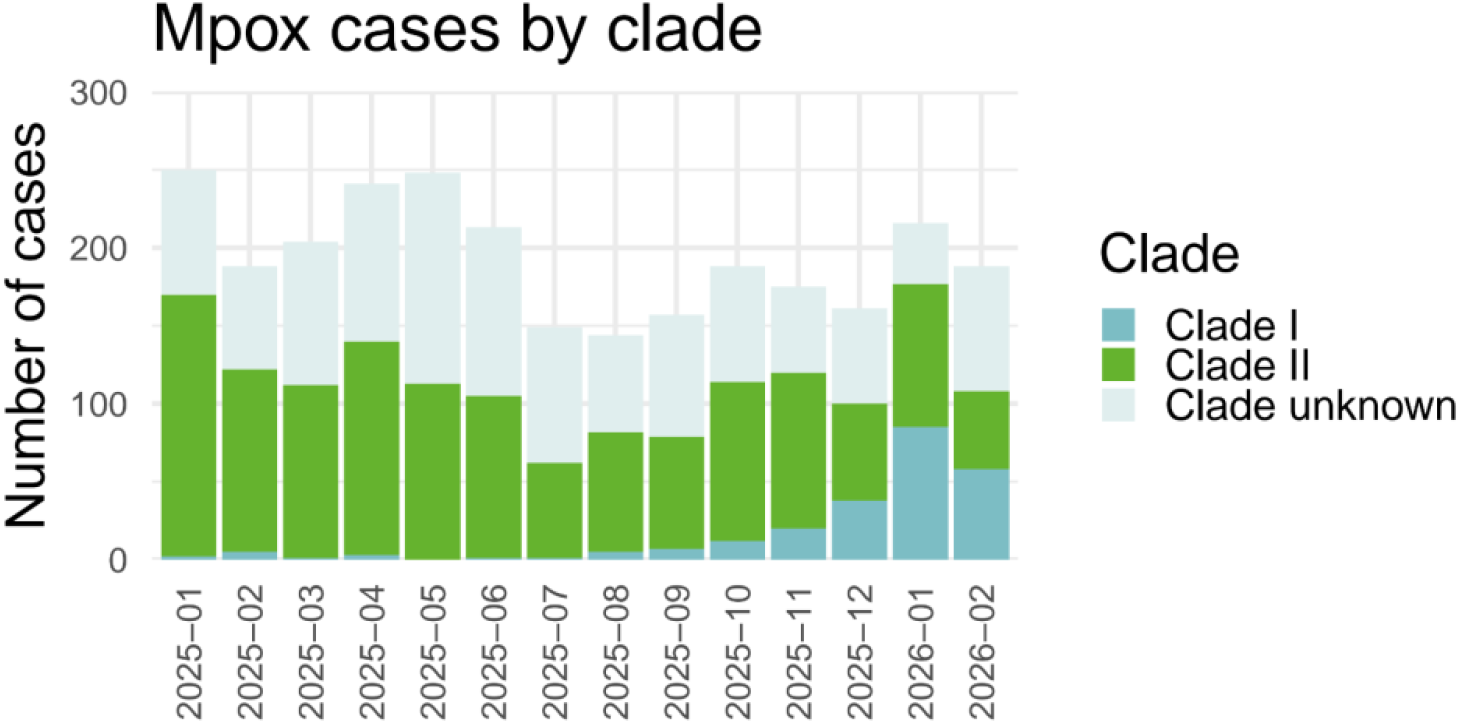
The proportion of mpox cases in the EU/EEA due to MPXV clade I and clade II. The data was extracted from The European Surveillance System (TESSy) on 9 March 2026.

While there is a lack of evidence on the vaccine effectiveness for MPXV clade I, around 69% of MPXV clade I cases among MSM in the EU/EEA were among unvaccinated individuals [9]. This indicates that there is an unrealised benefit of increasing the vaccine uptake among risk groups, even though the specific vaccine effectiveness against clade I is unknown.

The focus of our work is to assess the potential of additional vaccination campaigns among MSM, and our contribution is twofold. First, we estimate the vaccine uptake among MSM, by using key data from European MSM and Trans Persons Internet Survey 2024 (EMIS-2024), which included key mpox-specific questions (e.g., mpox vaccination and recovery) [10, 11]. Specifically, we provide three estimates for 28 of the 30 countries in the EU/EEA (excluding Liechtenstein and Iceland due to sample sizes below 100 respondents): i) the current vaccine uptake among MSM; ii) the hypothetical uptake when vaccinating MSM who would be willing to get vaccinated if mpox cases rise, and ; iii) the hypothetical uptake when vaccinating MSM who were willing to get vaccinated but had no vaccines available to them. Furthermore, we estimate the proportion of MSM that are recovered from mpox. Second, building on the three vaccine uptake estimates, we quantify the potential of increasing the vaccine uptake on the transmission of mpox among MSM, specifically, the reduction of the effective reproduction number.

## Methods

### Estimating the mpox vaccine uptake and natural immunity among MSM in the EU/EEA

We estimate the level of natural immunity and the vaccine uptake among MSM – both the current uptake and the uptake that could be attained by vaccinating every individual who is, or was, willing to get vaccinated by the time of responding to EMIS-2024. To estimate these quantities, we rely on three questions of EMIS-2024 [11, 12]. Specifically, we explore three vaccination scenarios, see Table 1: The baseline vaccination scenario is an estimate of the current vaccine uptake; the increased vaccination scenario A is an estimate of the hypothetical uptake that could be achieved if mpox cases rise; and the increased vaccination scenario B is an estimate of the hypothetical uptake that could be achieved by addressing the willingness for vaccination that was unmet due to vaccine unavailability. We stress that scenarios A and B are an extension of the baseline scenario, i.e., everyone who is vaccinated in the baseline scenario is also vaccinated in scenarios A and B. The three scenarios only differ with respect to the two-dose vaccination uptake, since we make the conservative assumption that the one-dose uptake is the same across all scenarios.

**Table 1.** Vaccination scenarios. The three considered scenarios of mpox vaccination, which are based on EMIS-2024 questions Q417 (“Have you been vaccinated against Mpox (‘monkeypox’) since May 2022?”) and Q419 (“If there was a rapid increase in Mpox cases in your community, how likely would you […] Get vaccinated against Mpox”).

| Vaccination scenario | Individuals considered as vaccinated with one dose | Individuals considered as vaccinated with two doses |
| --- | --- | --- |
| **Baseline vaccination**<br>Estimate of current uptake | Replied to “Have you been vaccinated against Mpox (‘monkeypox’) since May 2022?”:<br>• “Yes – with one dose”<br>• “Yes – but I am not sure how many doses” | Replied to “Have you been vaccinated against Mpox (‘monkeypox’) since May 2022?”:<br>• “Yes – with two doses” |
| **Increased vaccination scenario A**<br>Hypothetical increased vaccine uptake, addressing the *future* intention to get vaccinated given rise in mpox cases. Indicative of the vaccine uptake that can be achieved in the future. | Replied to “Have you been vaccinated against Mpox (‘monkeypox’) since May 2022?”:<br>• “Yes – with one dose”<br>• “Yes – but I am not sure how many doses” | *Either* replied to “Have you been vaccinated against Mpox (‘monkeypox’) since May 2022?”:<br>• “Yes – with two doses”<br>*Or* replied to “If there was a rapid increase in Mpox cases in your community, how likely would you [...] Get vaccinated against Mpox”:<br>• “Strongly agree”<br>• “Agree” |
| **Increased vaccination scenario B**<br>Hypothetical increased vaccine uptake, addressing the *past* intention to get vaccinated. Indicative of the vaccine uptake that could have been achieved in the past. | Replied to “Have you been vaccinated against Mpox (‘monkeypox’) since May 2022?”:<br>• “Yes – with one dose”<br>• “Yes – but I am not sure how many doses” | Replied to “Have you been vaccinated against Mpox (‘monkeypox’) since May 2022?”:<br>• “Yes – with two doses”<br>• “No, I wanted to but it was not available to me” |

We explore two scenarios for the natural immunity against mpox among MSM, see Table 2: A baseline natural immunity scenario; and a high natural immunity scenario. In the high immunity scenario, there are more MSM considered as recovered from mpox than in the baseline scenario.

**Table 2.** Natural immunity scenarios. The two considered scenarios of recovery from mpox, which are based on EMIS-2024 question Q416 (“Have you had Mpox (‘monkeypox’) since 2022?”).

| Natural immunity scenario | Individuals considered as recovered |
| --- | --- |
| **Baseline natural immunity**<br>Conservative interpretation of replies. | Replied to “Have you had Mpox (‘monkeypox’) since 2022?”:<br>• “Yes, and I was diagnosed by a doctor”<br>• “Yes, but I was not diagnosed in a health care setting” |
| **High natural immunity**<br>Interpretation of replies implying more recovered MSM, likely yielding an upper bound. | Replied to “Have you had Mpox (‘monkeypox’) since 2022?”:<br>• “Yes, and I was diagnosed by a doctor”<br>• “Yes, but I was not diagnosed in a health care setting”<br>• “No, but I had symptoms which might have been caused by mpox” |

The number of sexual partners among MSM follows a heavy-tailed distribution [13-16]. Hence, it is key to incorporate the heterogeneous distribution of sexual partners and its association with the vaccine uptake and natural immunity, which we do in two steps, please see Appendix A for more details. First, we fit a discrete Weibull distribution to the number of non-steady partners over the last 12 months reported by EMIS-2024 (question Q183), truncated at 200 partners. As it is an uncertain parameter, we conduct a sensitivity analysis of increasing the truncation to 350 non-steady partners per year in Appendix B. Second, for each of the three vaccination scenarios in Table 1 and each of the two natural immunity scenarios in Table 2, we fit Bayesian mixed-effects logistic regression model (BME) using the R package brms [17], an interface to Stan [18], for the probability of an individual to be vaccinated and recovered, respectively, depending on the number of non-steady partners and the country.

### Modelling the potential of future vaccination campaigns on reducing mpox transmission among MSM

Following the introduction of mpox into an MSM network, the outbreak dynamics crucially depend on the effective reproduction number ℛ_eff_, which equals the expected number of secondary cases produced by a typical^1^ infected individual and is a crucial threshold parameter [19-21]: If ℛ_eff_ ≤ 1, there is no major outbreak and the epidemic dies out rapidly, with only sporadic cases occurring. In contrast, ℛ_eff_ > 1 implies a non-zero probability for a major outbreak, and the larger ℛ_eff_, the larger the major outbreak probability and the expected outbreak size (all else being the same). In the following, our focus is therefore to estimate the effective reproduction number ℛ_eff_ to assess the effect of increasing the mpox vaccination uptake among MSM. We set the vaccine effectiveness against infection to VE_2_ = 82% and VE_1_ = 72% for two-dose and one-dose vaccination, respectively [22].

To compute the effective reproduction number ℛ_eff_, we describe the spread of mpox among MSM by a susceptible-infectious-recovered (SIR) model on a sexual contact network, analogously to Ball and Neal [23]. Susceptible (healthy) individuals can become infected upon contact with infectious individuals. Infectious individuals spread mpox to their contacts until they recover (cure) according to an exponential distribution with an average duration of 1/*δ* = 10.5 days [24]. Individuals in the recovered compartment are immune against mpox due to natural and/or vaccine-induced immunity.

We formulate a sexual contact network to capture the heterogeneous, heavy-tailed distribution of the number of sexual partners in MSM networks [13] as well as different types of sexual partnerships (steady or non-steady). Specifically, we use a two-layered configuration model network as in [25], where layer *l* = 1 captures steady contacts and layer *l* = 2 captures non-steady contacts. The random variable *D*_*l*_ denotes the number of susceptible sexual partners (degree) on layer *l* of a susceptible individual in the baseline vaccination scenario in Table 1, which is informed by the fitted BMEs of the recovery status and vaccination status and EMIS-2024 data, see Appendix C. We denote the infection rate per sexual partner on layer *l* by *β*_*l*_ = *αc*_*l*_, where *α* denotes the infection rate multiplier. Furthermore, *c*_1_ = 34.10/365 and *c*_2_ = 3.23/365 denote the relative rate of contacts per partner on layers 1 and 2, obtained from fitting to the EMIS-2017 data on the recency of sex [25]. This corresponds to an average of 34.10 sexual contacts per year within steady partnerships (layer 1) and 3.23 contacts per year within casual partnerships (layer 2).

After the introduction of an infection into an MSM network of sufficient size, the early phase of the outbreak is approximated by a branching process [23, 26, 27]. The early phase of the outbreak crucially depends on the effective reproduction number ℛ_eff_, which equals the dominant eigenvalue *λ*_1_ of the 2 × 2 mean offspring matrix

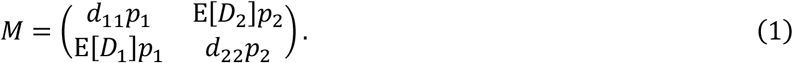

Here, the probability of transmission from an infectious individual to a susceptible individual given a sexual contact on layer *l* equals

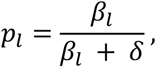

and the expected excess degree of layer *l* equals

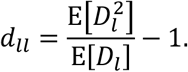

Starting from the baseline vaccination scenario in Table 1, we calibrate the SIR model to the effective reproduction number ℛ_eff_, which varies substantially over time and across countries in the EU/EEA, reflecting time-varying behaviour and regional differences in sexual behaviour, surveillance, and population-level immunity. To capture these variations, we explore a wide range of values from ℛ_eff_ = 1 to ℛ_eff_ = 4, which covers the span of the reproduction number estimated from mpox case data in the EU/EEA over the last 12 months, see Appendix D. For each of these values for ℛ_eff_, we calibrate the model by setting the infection rate multiplier *α* (using bisection), which relates to the effective reproduction number ℛ_eff_ via a one-to-one mapping.

In contrast to the *basic* reproduction number ℛ_0_ that assumes a naïve population, the effective reproduction number ℛ_eff_ does not assume that everyone is susceptible and therefore depends on the number of vaccinated and recovered individuals. Both of the increased vaccination scenarios A and B decrease the number of susceptible individuals, which translates to fewer susceptible contacts in both layers of the sexual contact network. Furthermore, recovery from mpox reduces the number of susceptible individuals, and we assume that, conditional on the number of non-steady sexual partners, the recovery and vaccination status is stochastically independent. For the increased vaccination scenarios A and B, we denote the number of susceptible sexual partners (the degree distribution) on layer *l* by 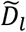 and the resulting effective reproduction number by 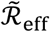, which follows from (1) using the same probability *α* as for the baseline vaccination scenario. Then, as the key metric to quantify the effect of increasing the vaccine uptake, we focus on the relative reduction^2^ 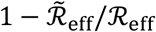.

We inform the model by parameters of MPXV clade II, for the lack of MPXV clade I parameters in the EU/EEA context. To address the impact of uncertain clade I parameters, we explore scenarios of reduced vaccine effectiveness against clade I and/or increased transmissibility of clade I in Appendix E. We stress that Appendix E therefore addresses the important interaction of possibly reduced but uncertain vaccine effectiveness (via different scenarios) and uncertain vaccine uptake (captured by the Bayesian approach).

## Results

Figure 2 shows the estimated mpox vaccine uptake among MSM for the three vaccination scenarios in Table 1. We stress four observations. First, the estimated baseline vaccine uptake varies strongly across countries, ranging from country-specific medians of 1.0% to 31.4%. Particularly, the results suggest that few MSM have been vaccinated against mpox in some countries. Second, for all countries the potential uptake in the increased vaccination scenario A is substantially higher than the estimated current vaccine uptake (baseline), quantified by the respective median uptake of the scenarios across countries of 75.0% versus 13.6%. Hence, the results suggest a large potential willingness for mpox vaccination among MSM across EU/EEA, if the epidemic situation deteriorates (cases rise). Third, for most countries, the potential vaccine uptake in scenario B is substantially larger than for the baseline scenario (median across countries of 32.8% versus 13.6%), which indicates a large unmet willingness for mpox vaccination among MSM since 2022. Fourth, the potential vaccine uptake is larger for scenario A than for scenario B, which shows that the willingness to get vaccinated if cases rise is substantially larger than the past unmet willingness for vaccination. Lastly, we note that due to a limited sample size for some countries (e.g. for Luxembourg), the posterior estimate for the vaccine uptake is associated with large uncertainties. For the values underlying Figure 2, we refer to Table A7 in Appendix A.

**Figure 2.**
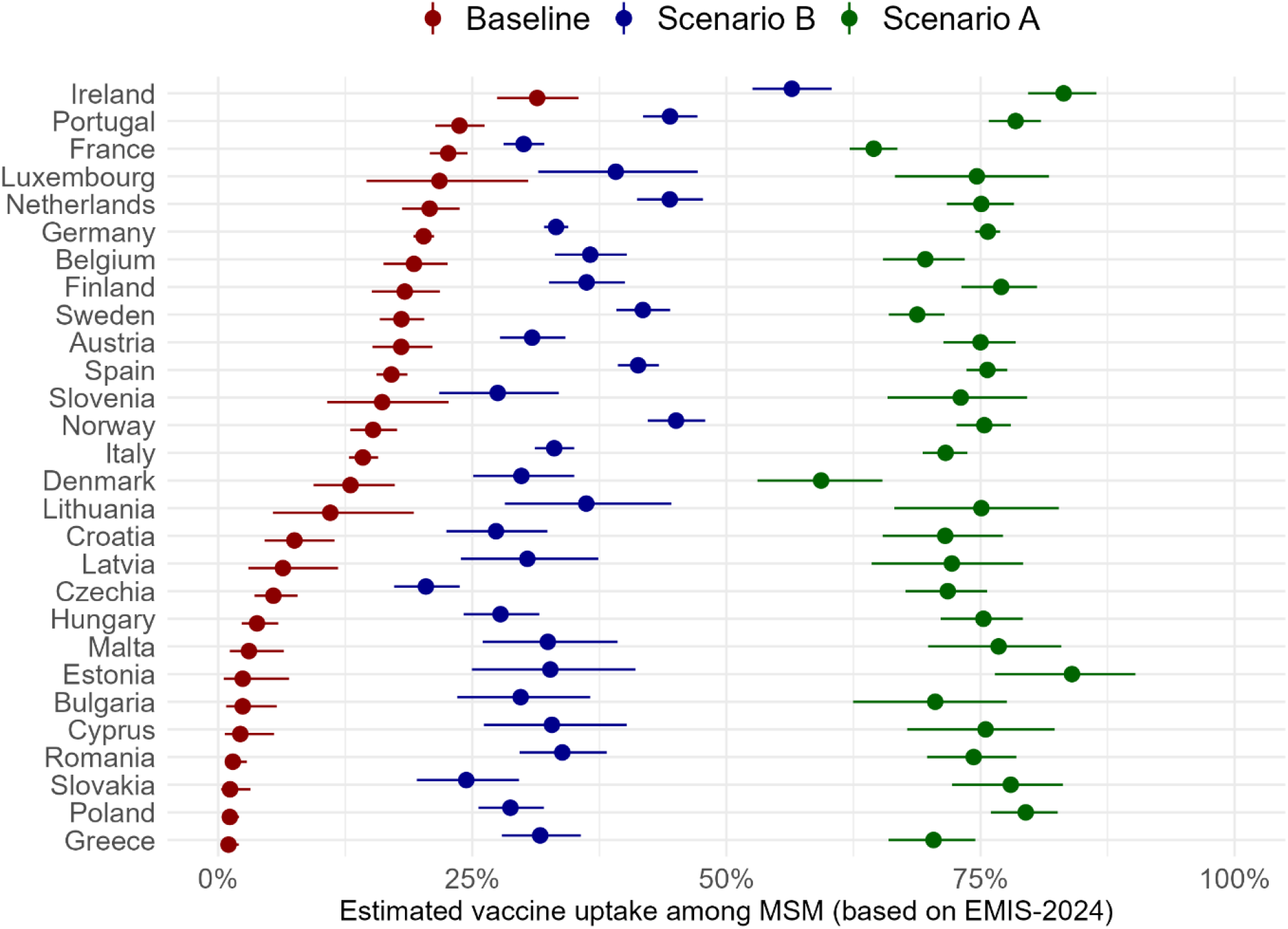
Estimated mpox vaccine uptake among MSM in the EU/EEA. The three scenarios show the estimates for the two-dose uptake corresponding to: The current vaccine uptake among MSM (“Baseline”); the current vaccine uptake plus vaccinating all MSM who would like to receive vaccinations given a rapid rise in mpox cases (“Scenario A”); and the current vaccine uptake plus vaccinating all MSM with a past willingness to get vaccinated (“Scenario B”). The points correspond to the respective median, and the width of the lines shows the 95% uncertainty interval, obtained from 100 posterior samples of the Bayesian mixed-effects logistic regression model of the vaccine status versus the number of sexual partners fitted to EMIS-2024 data.

Figure 3 shows the estimated proportion of MSM that are recovered from mpox for the two natural immunity scenarios in Table 2. We make four observations. First, the estimated proportion of natural immunity is relatively low as the median estimate does not exceed 5% in any of the countries or scenarios. While the high natural immunity scenario does include self-diagnosed cases with possibly mild symptoms (participants replying “No, but I had symptoms which *might have been* caused by mpox”) and therefore addresses some extent of underreporting, mpox infections that are completely asymptomatic are not captured, which might result in an underestimation of the level of natural immunity in the presence of many asymptomatic infections. Second, the probability of having recovered from mpox is markedly higher for individuals with many sexual partners, see Figures A5 and A6 in Appendix A for the detailed dependency of recovery status on the number of partners. Third, the estimated level of natural immunity varies markedly across countries, ranging from 0.75% for Romania to 3.07% for Spain (median estimates in the baseline scenario). Fourth, while the estimated natural immunity levels do depend on the baseline and high natural immunity scenarios, the extent is not very large (median estimate of 1.25% versus 1.79%) and the uncertainty intervals are overlapping for most countries. As for the estimated vaccine uptake, we note that due to a limited sample size, there are large uncertainties in the estimates for the level of natural immunity for some countries. For the values underlying Figure 3, we refer to Table A8 in Appendix A.

**Figure 3.**
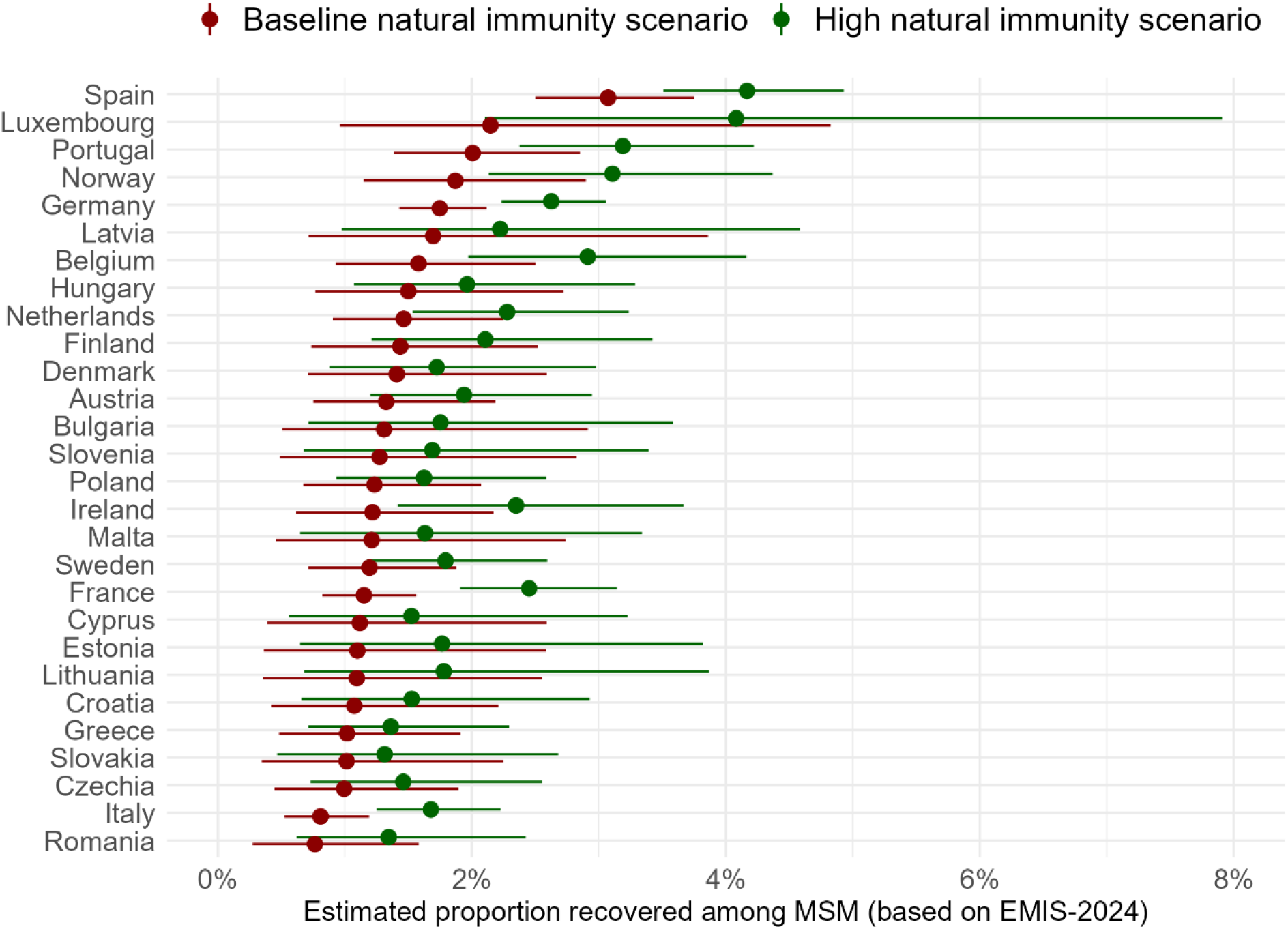
Estimated proportion of MSM recovered from mpox in the EU/EEA. The two scenarios show the natural immunity estimates corresponding to: A more conservative interpretation of EMIS-2024 responses (“Baseline natural immunity scenario”); and an alternative interpretation of EMIS-2024 responses leading to more recovered MSM (“High natural immunity scenario”). The points correspond to the respective median, and the width of the lines shows the 95% uncertainty interval, obtained from 100 posterior samples of the Bayesian mixed-effects logistic regression model of the mpox recovery status versus the number of sexual partners fitted to EMIS-2024 data.

Figure 4 shows the estimated effect of increasing the vaccine uptake among MSM for vaccination scenarios A and B. While there are some variations across countries, the estimated effect is consistently high: Compared to the baseline vaccination scenario, the effective reproduction number ℛ_eff_ decreases by a median of 57.1% (ranging from 37.6-74.8% across countries) in vaccination scenario A and by 32.6% (ranging from 8.5-42.9% across countries) in vaccination scenario B.

**Figure 4.**
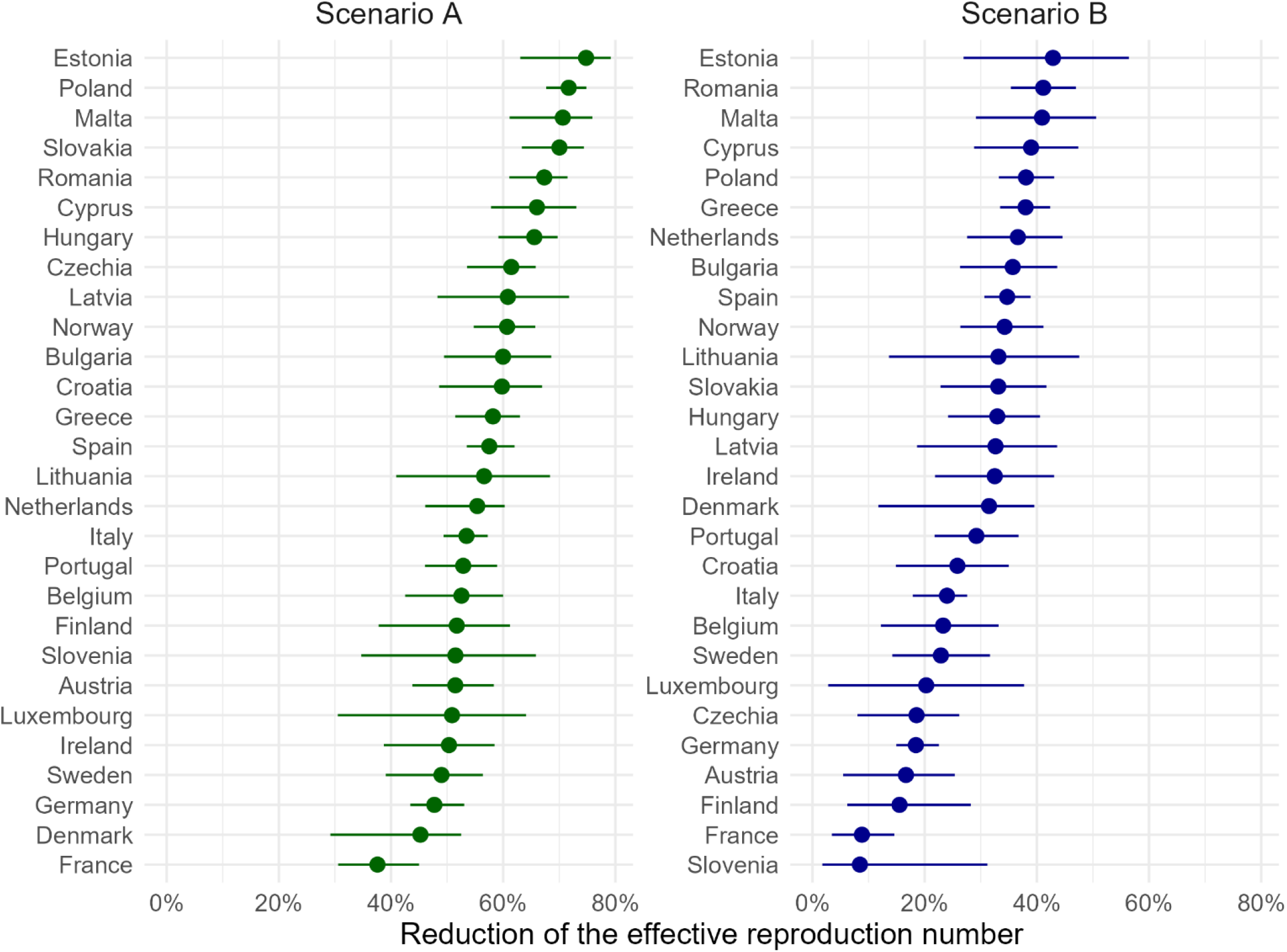
The potential of increasing the mpox vaccine uptake among MSM to reduce transmission. The reduction of the effective reproduction number ℛ_eff_, obtained by comparing the increased vaccination scenarios A and B with the baseline vaccination scenario. The results are aggregated over the baseline and high natural immunity scenarios as well as aggregated over a range of the effective reproduction number from ℛ_eff_ = 1 to ℛ_eff_ = 4 in the baseline vaccination scenario. The points correspond to the respective median, and the width of the lines shows the 95% uncertainty interval, obtained from 100 posterior samples of the Bayesian mixed-effects logistic regression models of the vaccine and recovery status versus country and number of sexual partners fitted to EMIS-2024 data.

The results in Figure 4 apply to MPXV clade II and provided similar parameters of the two clades, also to clade I. However, since the parameters of clade I are uncertain, we explore further scenarios in Appendix E. Specifically, we explore the impact of a higher transmissibility of clade I versus clade II and/or the impact of reduction of the vaccine effectiveness against infection, which could be due to, e.g., waning of vaccine-induced immunity or a lower vaccine effectiveness against clade Ib as compared to clade II.

## Discussion

The contributions of this study are threefold. First, we estimate the current mpox vaccine uptake among MSM for 28 of the 30 countries in the EU/EEA by using EMIS-2024 data (“Baseline vaccination scenario”). Estimates of the vaccine uptake are crucial for assessing current preparedness levels and identifying gaps across the EU/EEA. We note that our uptake estimates are the first that cover EU/EEA countries using data from a study sample; whereas it is harder to obtain the vaccine uptake from mpox dose administration data due to the uncertain denominator (i.e., the MSM population size) as done, e.g., in [25]. Our results show that there are substantial differences of the vaccine uptake across the EU/EEA. Thus, for assessing response and preparedness measures against mpox, it is crucial to take into account the specifics of the respective country. Despite variations across countries, our results suggest a substantial potential for increasing the vaccine uptake among MSM by future campaigns – provided that the epidemiological situation deteriorates and mpox cases rise (scenario A). In agreement with this future intention to get vaccinated, our results suggest that there was a substantial unmet willingness for mpox vaccination among MSM since 2022 (scenario B). Nevertheless, it is important to underscore that access to mpox vaccines within EU/EEA countries has undergone substantial changes between 2022 and the present. In 2022, limited vaccine availability across the EU/EEA constrained the capacity to meet vaccination demand, leading national authorities to implement prioritisation strategies to available doses and hence, it is possible that some of the EMIS-2024 respondents had no opportunity to receive mpox vaccination despite their willingness.

Second, we provide estimates for the levels of natural immunity against mpox among MSM for 28 countries in the EU/EEA. Our results suggest that there is a relatively low proportion of recovered individuals (less than 5% of MSM for any country). For the baseline and high natural immunity scenarios, the median estimate across countries is 1.25% and 1.79%, respectively (see Table A8). These EMIS-2024 based estimates are in good agreement with estimates based on The European Surveillance System (TESSy) data, which resulted in a proportion of 1.33% of MSM recovered from mpox in 2022 by assuming that 2.8% of men are MSM [25]. Furthermore, our results suggest that the recovery against mpox is mostly concentrated among MSM with many sexual partners (Figures A5 and A6). This implies that the protective effect of accumulated natural immunity could decrease substantially if the composition of susceptible MSM with many partners changes in the future, for instance, due to new young people entering MSM sexual networks, or due to behavioural change of some individuals who change from fewer to more sexual partners. Underlying the aggregate estimates for the vaccine uptake and natural immunity, our results provide an estimate for the individual vaccine and recovery status disaggregated by the number of sexual partners, which is crucial to parametrising mathematical models of mpox transmission.

Third, utilising the results on vaccine-induced and naturally-acquired immunity, we develop a mathematical model to estimate the effect of increasing the vaccine uptake among MSM by targeting individuals that are willing to get vaccinated. We estimate a substantial reduction of the effective reproduction number (Figure 4), which suggests that many outbreaks among MSM can be (scenario A) or could have been (scenario B) prevented by increased vaccination. The results in Figure 4 are based on vaccination scenarios A and B in Table 1, which consider that *all* MSM are vaccinated who are/were willing to. While these scenarios are insightful to obtain a reasonable best-case potential of vaccination campaigns, administering mpox vaccines is subject to practical challenges, including reaching individuals at risk, availability of vaccines (particularly in remote regions), and situational awareness of target groups. Consequently, it may be difficult to achieve the vaccine uptake given by scenarios A and B. Nonetheless, as shown by Figure A13 in Appendix E, our results suggest a large potential even if the vaccine reaches only a proportion of individuals who are willing to get vaccinated.

In summary, this work suggests not only a substantial unmet willingness of MSM to get vaccinated against mpox, but also a large potential of increasing the vaccine uptake in reducing transmission of mpox in MSM sexual networks in the EU/EEA. Increasing vaccine uptake among MSM depends on making the vaccine accessible to the population, both in terms of cost as well as logistics. Additionally, good collaboration with civil society organisations that work for MSM is essential to raise awareness of the need to get vaccinated and thus increase the vaccine uptake.

### Limitations

This study is subject to several limitations. First and foremost, EMIS-2024 employed non-representative online convenience sampling. It is plausible that the respondents have more sexual partners than MSM who did not participate in the survey. If true, it limits the ability to generalise the results to all MSM, however given that the 2022 mpox outbreak disproportionately affected MSM with large numbers of partners, we believe that the results of this work are key to understanding the impact of prevention efforts. However, we emphasise that the estimates for the vaccine uptake among MSM should be interpreted with caution, and it remains a crucial open question to which extent the estimates hold for the general MSM population. Furthermore, the willingness to get vaccinated against mpox might have changed since the data collection of EMIS-2024, for instance, due to a decreasing risk perception.

Furthermore, the mathematical model that we used to assess the effect of vaccination on the reproduction number is, as any model, affected by the uncertainty of mpox parameters. Specifically, lacking evidence, we assume that MPXV clade I and clade II have a similar transmissibility and that the vaccine effectiveness is the same for both clades. As these two parameters seem crucial for assessing the effect of vaccination, we provide a sensitivity analysis in Appendix E, see Figures A11 and A12. These sensitivity analyses also capture a reduction of the vaccine effectiveness due waning over time. Furthermore, we assume that individuals that are recovered from mpox are protected from reinfections, hence, our results hold as long as the proportion of infections seen among previously infected people is negligible. We assumed that previous clade II infection confers the same immunity against clade I infections as against clade II infections. However, we note that, if there is evidence that points towards significant levels of reinfections, then our methodology can be adjusted to include reinfections.

Lastly, our work focuses solely on the spread of mpox among MSM, since this group has been disproportionately affected by mpox outbreaks in the EU/EEA. If future evidence shows that population groups other than MSM are significantly affected by mpox in the EU/EEA, our results on the reproduction number need to be revised.

## Data Availability

All data produced in the present study are available upon reasonable request to the authors.

## Author contribution statements

BP: Conceptualisation, Data curation, Formal analysis, Methodology, Software, Visualisation, Writing – original draft, Writing – review & editing; DH: Formal analysis, Methodology, Writing – review & editing; LA, KJJ: Investigation, Data curation, Writing – review & editing; JBP, XA: Investigation, Validation, Writing – review & editing; AP, DP: Supervision, Writing – review & editing; AJS: Investigation, Data curation, Writing – review & editing; LN: Investigation, Supervision, Writing – review & editing

## Acknowledgements

We begin by thanking everybody who took part in EMIS-2024.

## EMIS-2024 Execution

EMIS-2024 is executed by a consortium of three partners: Deutsche Aidshilfe (DAH), Maastricht University, and Robert Koch Institute (RKI). Team DAH: Dr. Axel J. Schmidt (co-principal investigator), Dr. Tamás Bereczky (coordinator EEA). Team Maastricht: Prof. Dr. Kai J. Jonas (co-principal investigator), Liana Aphami (data coordinator), Jules L. Casalini (trans sub-survey). Team RKI: Dr. Ulrich Marcus (co-principal investigator), Dr. Nikolay Lunchenkov (coordinator EECA).

## EMIS-2024 Funding

German Ministry of Health (*Bundesministerium für Gesundheit*, **DE**) / Global Health Protection Programme (GHPP, **DE**); European Centre for Disease Prevention and Control (ECDC, **EU**); Swiss AIDS Federation, Swiss Federal Office of Public Health (**CH**); Luxembourg Health Directorate (**LU**); Norwegian Directorate of Health (*Helsedirektoratet*, **NO**); The Public Health Agency of Sweden (*Folkhälsomyndigheten*, **SE**); Sciensano (*Institut Scientifique de Santé Publique*, **BE**); Estonian Ministry of Social Affairs (*Sotsiaalministeerium*, **EE**); Health Service Executive (**IE**); Portuguese Directorate-General of Health (*Direção-Geral da Saúde*, **PT**); Spanish Ministry of Health (*Ministerio de Sanidad*, **ES**); Central Health Department, Israeli Ministry of Health (הבריאות משרד, **IL**); National Institute for Public Health and the Environment (RIVM) / *SOAids Nederland* (**NL**).

The following list acknowledges all partners in EMIS by country covered in this manuscript. Individual names are mentioned if a freelancer was the main contact and/or translator or where input on the questionnaire development came from a person not formally representing an organisation. The order (if available) is: main NGO partner, other NGO partners, academic partners, governmental partners, individuals.

## Europe

Grindr & Grindr for Equality, ROMEO, Hornet.

## AT

*Aids Hilfe Wien*. **BE**: Sensoa, Exaequo, Sciensano. **BG**: GLAS Foundation. **CY**: AIDS Solidarity Movement, Christos Krasidis. **CZ**: *Česká společnost AIDS pomoc*. **DE**: *Deutsche AIDS-Hilfe*, Robert Koch Institute, IWWIT. **DK**: *Statens Serum Institut*, Dr. Susan Cowan, Dr. Maria Wessmann. **EE**: Estonia National Institute for Health Development, Dr. Kristi Rüütel, Dr. Sigrid Vorobjov. **ES**: SEISIDA, STOPSIDA, CEEISCAT, *Ministerio de Sanidad*. **FI**: *Positiiviset ry*. **GR**: Positive Voice. **HR**: Iskorak, Zoran Dominković. **HU**: Háttér Society. **IE**: EMIS-2024 Ireland Promotion Sub-Committee, Health Service Executive, Mick Quinlan. **IT**: Fondazione LILA Milano ONLUS, University of Verona. **LT**: Demetra, National Public Health Center. **LV**: AGIHAS. **LU**: *Luxembourg Health Directorate*. **MT**: HIV Malta, Infectious Disease Prevention and Control Unit. **NL**: SOAids, Maastricht University, RIVM; **NO**: Norwegian Institute of Public Health, Dr. Rigmor C. Berg. **PL**: *Fundacja Edukacji Społecznej*. **PT**: GAT Portugal, *Instituto de Saúde Pública da Universidade do Porto*, Portuguese Directorate-General of Health. **RO**: ARAS, Tudor Kovacs. **SE**: RFSL, *Folkhälsomyndigheten*. **SI**: LEGEBITRA, ŠKUC. **SK**: Pride Kosice.

## Other

Sigma Research, London School of Hygiene & Tropical Medicine.

## Appendix A. Estimating the vaccine-induced and naturally acquired immunity against mpox among MSM in the EU/EEA

### The distribution of non-steady sex partners

To obtain the distribution of non-steady (casual) sex partners, we rely on EMIS-2024 question Q183, which reads “How many different non-steady male partners have you had sex with in the last 12 months?” Respondents could reply with the exact number of partners if it did not exceed 10, or reply by stating the range of “11-20”, “21-30”, “31-40”, “41-50”, or “more than 50”. Using the aggregated data from all countries (acknowledging that fitting a heavy tailed distribution requires many samples), we fit a discrete Weibull distribution, see the pdweibull function of the extraDistr package [28], truncated at 200 non-steady partners by maximising the likelihood. The truncation at 200 partners is in line with the National Surveys of Sexual Attitudes and Lifestyles (Natsal) from the UK; In Natsal-2 and Natsal-3, a maximum of 250 and 100 partners per year, respectively, was reported [14, 29]. However, the maximum number of partners in a given MSM sexual network is uncertain and difficult to obtain. Certainly, higher values for the number of partners have been reported, see for example the work by Mendez-Lopez et al. [16] that uses thirteen national and multi-national surveys, which suggests annual non-steady partner numbers exceeding 200. While we consider 200 non-steady partners as an upper bound for many MSM networks, we acknowledge that higher values are possible for some networks. Hence, to address MSM networks whose maximum number of non-steady partners is higher than 200, we provide a sensitivity analysis in Appendix B that considers 350 non-steady partners. We emphasise that our method can use any maximum number of non-steady partners provided.

The parameters that maximised the likelihood (for 200 partners) were shape1=0.791 and shape2=0.631. Figure A1 shows that the discrete Weibull distribution fits the EMIS-2024 data well.

**Figure A1.**
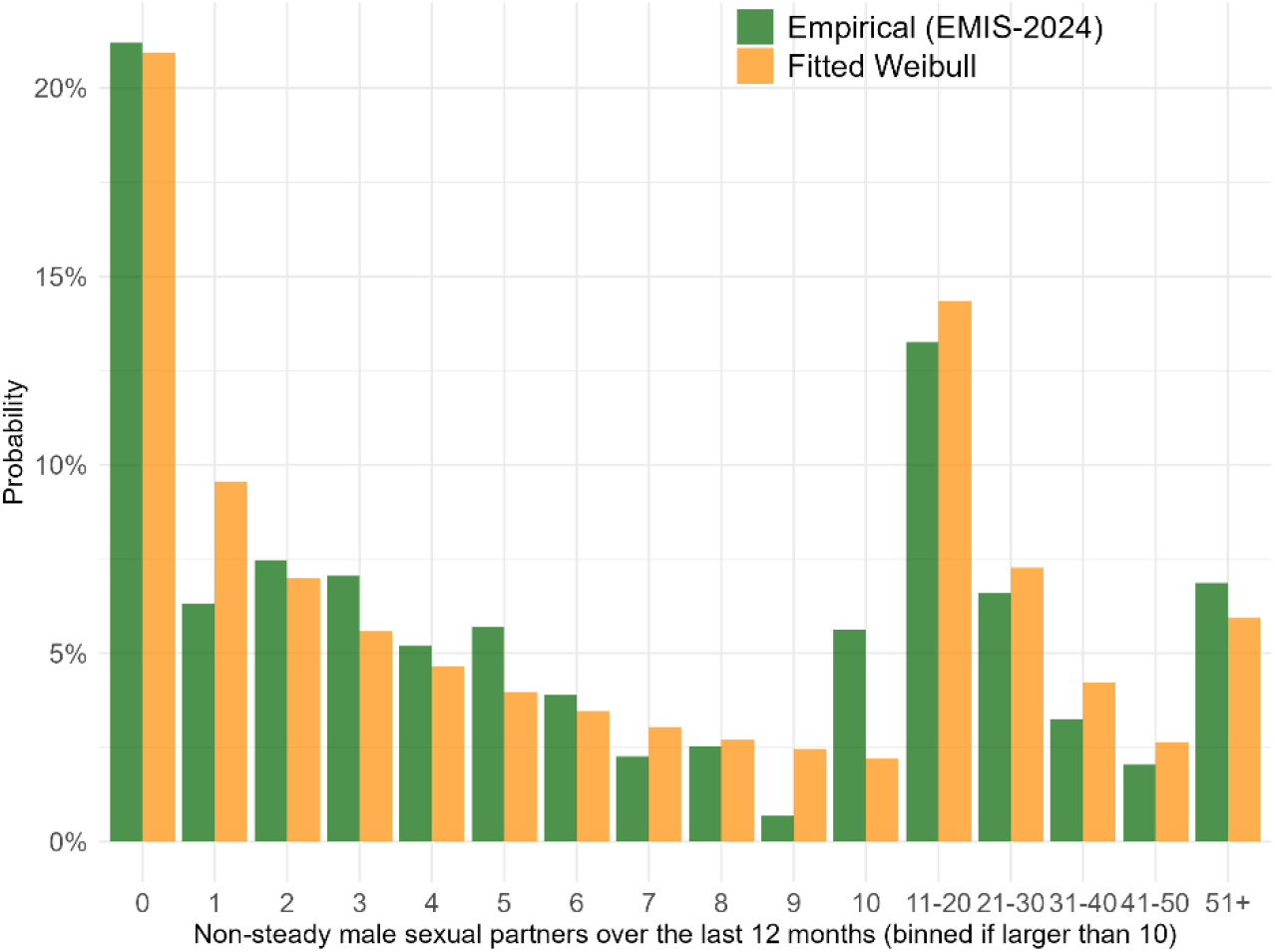
Fitting to the number of non-steady sexual partners.

### The distributions of vaccination and recovery status

In the following, we detail our approach of estimating the probability of an MSM individual to be vaccinated against mpox and/or recovered from mpox. To do this, we fit several instances (six instances in total) of a Bayesian mixed-effects logistic regression model (BME). Each instance of the model corresponds to a distinct binary outcome *X*_*i*_ and probability *p*_*i*_, defined further below. For each instance, the outcome *X*_*i*_ = 1 occurs with probability *p*_*i*_ for an individual *i* in country *c*(*i*) with *d*_*i*_ non-steady sexual partners over the last 12 months. Hence, the outcome *X*_*i*_ = 0 occurs with probability 1 − *p*_*i*_. The six instances of the model are given by:

- Three model instances for the probability of being vaccinated with two doses for each of the three vaccination scenarios in Table 1. In this case, *X*_*i*_ = 1 denotes that individual *i* has received two doses, and *X*_*i*_ = 0 indicates that individual *i* has not received two doses. Hence, if *X*_*i*_ = 0, then individual has received zero doses or one dose. To be precise, we fit one joint model with the scenario as a variable (via the formula “0 + scenario + logx:scenario + (0 + scenario + logx:scenario | country)” in R, where logx corresponds to log (1 + *d*_*i*_)) and obtain the three models for each vaccination scenario by setting the scenario variable to the respective vaccination scenario.
- One model instance for the *conditional* probability of being vaccinated with one dose, given that individual *i* has not received two doses (i.e., given that individual *i* is unvaccinated or has received one dose). Here, *X*_*i*_ = 1 denotes that individual *i* has received one dose, and *X*_*i*_ = 0 means that individual *i* is unvaccinated. Then, the unconditional probability for being vaccinated with one dose follows with the two-dose model for the baseline vaccination scenario. By choosing to fit the conditional probability, we avoid the issue that the unconditional probabilities for zero, one, and two doses exceed 1. Since the one-dose uptake does not change across the scenarios, there is only one model for the one-dose vaccination uptake.
- Two model instances for the probability of being recovered from mpox for each of the two recovery scenarios in Table 2. Here, *X*_*i*_ = 1 denotes that individual *i* has recovered from mpox, and *X*_*i*_ = 0 that individual *i* has not had mpox.

In any of the six model instances, the dependency of the probability *p*_*i*_ on the number of partners *d*_*i*_ and the country *c*(*i*) is modelled as

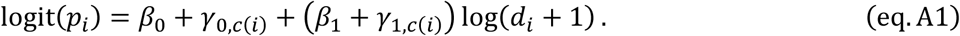

Here, the logit function is given by 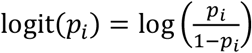. The coefficients *β*_0_ and *γ*_0,*c*(*i*)_ are the fixed effect intercept and the country-level intercept, and the coefficients *β*_1_ and *γ*_1,*c*(*i*)_ are the fixed intercept and the country-specific slopes. We used the default priors [17, 30] for the parameters, which are weakly informative. (A Gaussian distribution 𝒩(0,10) for fixed effects, Half-Student-t(3,0,10) distributions for the country-specific coefficients, and an LKJ(1) prior for group-level correlation matrices.) To translate the EMIS-2024 responses on the non-steady partners that are reported as intervals (“11-20”, etc.), we use the conditional mean of the fitted Weibull distribution given that the number of partners is in the respective interval.

The models (eq.A1) were fitted using Markov Chain Monte Carlo (MCMC) sampling via the brm command of the brms R package [17, 30], an interface to Stan [18], with 8 chains, 40,000 iterations per chain (including 20,000 warmup iterations), and the target average acceptance probability (“adapt_delta”) set to 0.95 to ensure adequate convergence. Model convergence was assessed using R-hat statistics, whose maximum was 1.00035 across all BMEs, and bulk effective sample sizes, whose minimum was 23628.84 across all BMEs. From the posterior distribution, we extracted 20,000 samples to quantify uncertainty in country-specific recovery rates across the range of sexual partner counts.

For the three respective vaccination scenarios in Table 1 (three scenarios for the two-dose uptake with the same one-dose uptake), Figures A2-A5 below show the results of fitting the BHMs (eq.A1) to EMIS-2024 data on the vaccination status. The resulting coefficients of the model (eq.A1) are stated in Tables A1-A4. Similarly, for two natural immunity scenarios in Table 2, Figures A6 and A7 below show the results of fitting the BHMs (eq.A1) to the EMIS-2024 data on the recovery status, with the resulting coefficients stated in Tables A5 and A6. The BHMs fit the data reasonably well. For some countries, the uncertainty intervals are rather wide due to a limited number of EMIS-2024 samples.

The proportions of vaccinated and recovered MSM, shown in Figures 1 and 2, are obtained by combining the fitted Weibull degree distribution with the posterior samples of the vaccination/recovery status conditional on the degree (law of total probability).

**Figure A2.**
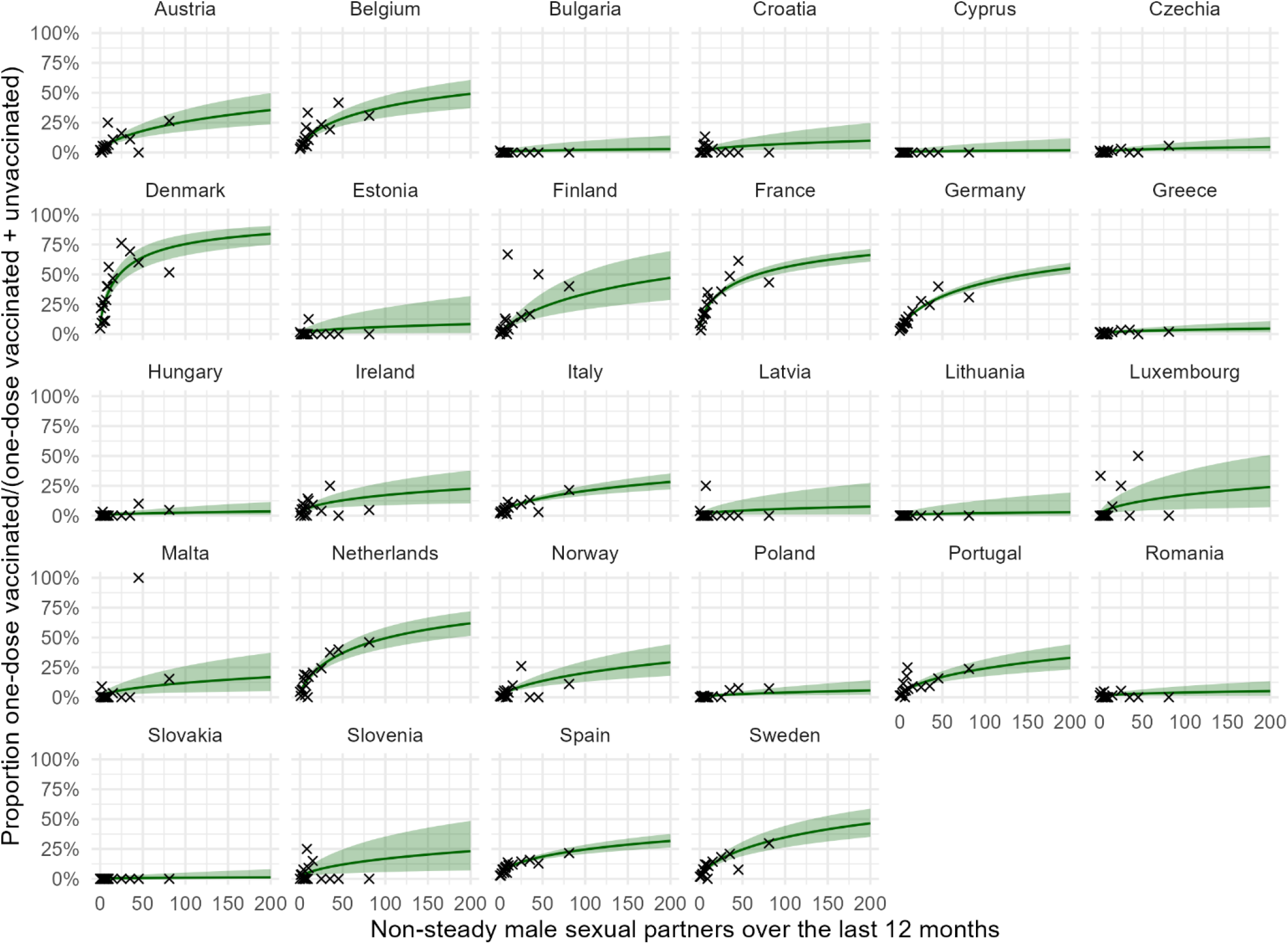
Estimated conditional one-dose vaccine uptake among MSM. The conditional probability of being vaccinated against mpox with one dose versus the number of non-steady male sexual partners over the last 12 months (or the degree), given on being vaccinated with at most one dose. The green line and green areas show the median and the 95% uncertainty interval of the posterior vaccination probability of the fitted BME (eq.A1), see Table A1 for more details on the posterior coefficients. The black crosses show the empirical fractions of vaccinated individuals by degree, where banded EMIS-2024 responses were mapped to numeric values using a Weibull degree distribution fitted to the data, providing a model-based alternative to the empirical approach described by Mendez-Lopez et al. [16]. Note that the denominator was small for some data points (which is not visible since the crosses are fractions), and therefore a worse fit to these data points did not influence the posterior as much as data points with a large denominator.

**Figure A3.**
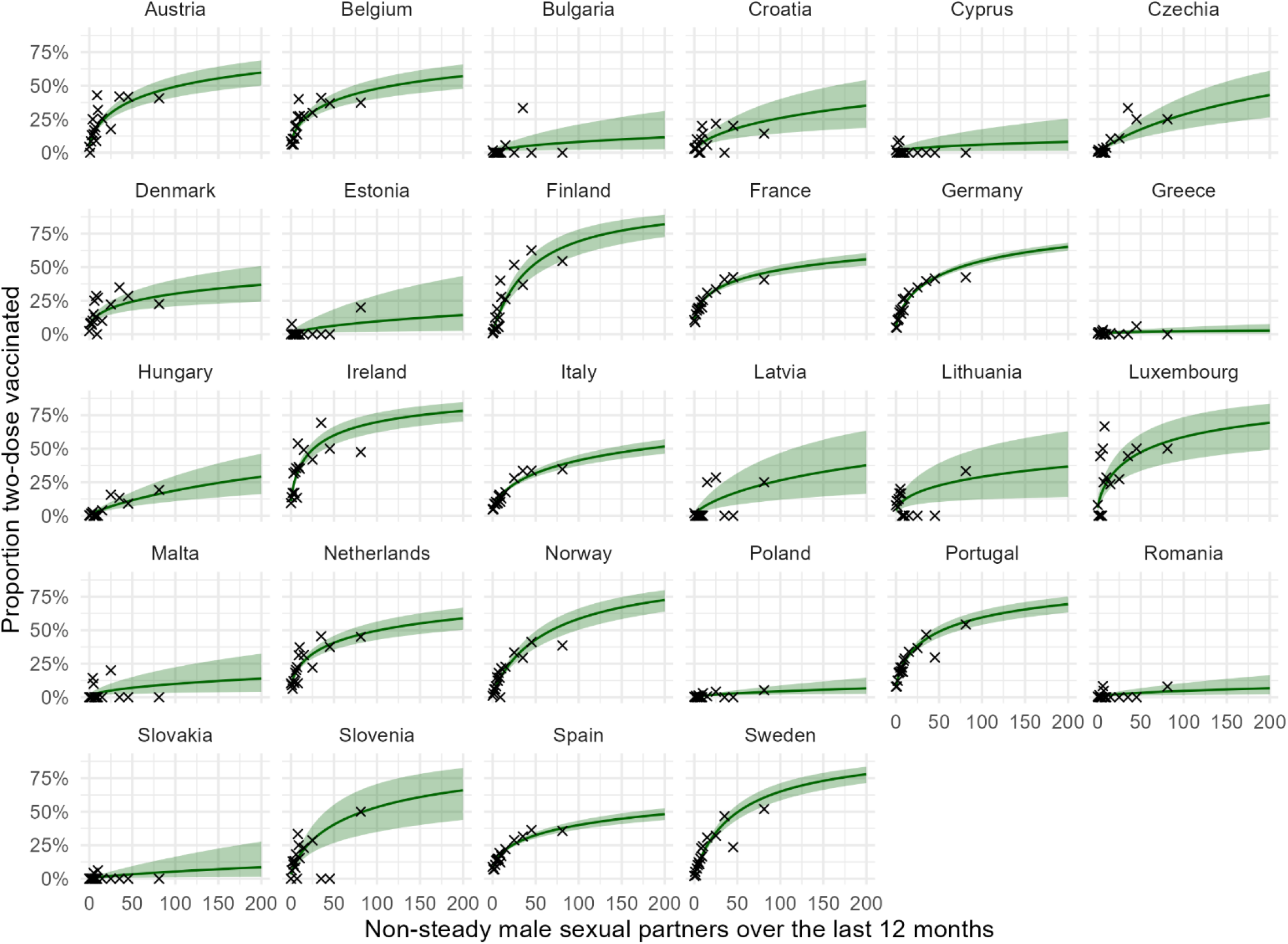
Estimated two-dose vaccine uptake among MSM (baseline vaccination scenario). The probability of being vaccinated against mpox versus the number of non-steady male sexual partners over the last 12 months (or the degree) for the baseline vaccination scenario. The green line and green areas show the median and the 95% uncertainty interval of the posterior vaccination probability of the fitted BME (eq.A1), see Table A2 for more details on the posterior coefficients. The black crosses show the empirical fractions of vaccinated individuals by degree, where banded EMIS-2024 responses were mapped to numeric values using a Weibull degree distribution fitted to the data, providing a model-based alternative to the empirical approach described by Mendez-Lopez et al. [16]. Note that the denominator was small for some data points (which is not visible since the crosses are fractions), and therefore a worse fit to these data points did not influence the posterior as much as data points with a large denominator.

**Figure A4.**
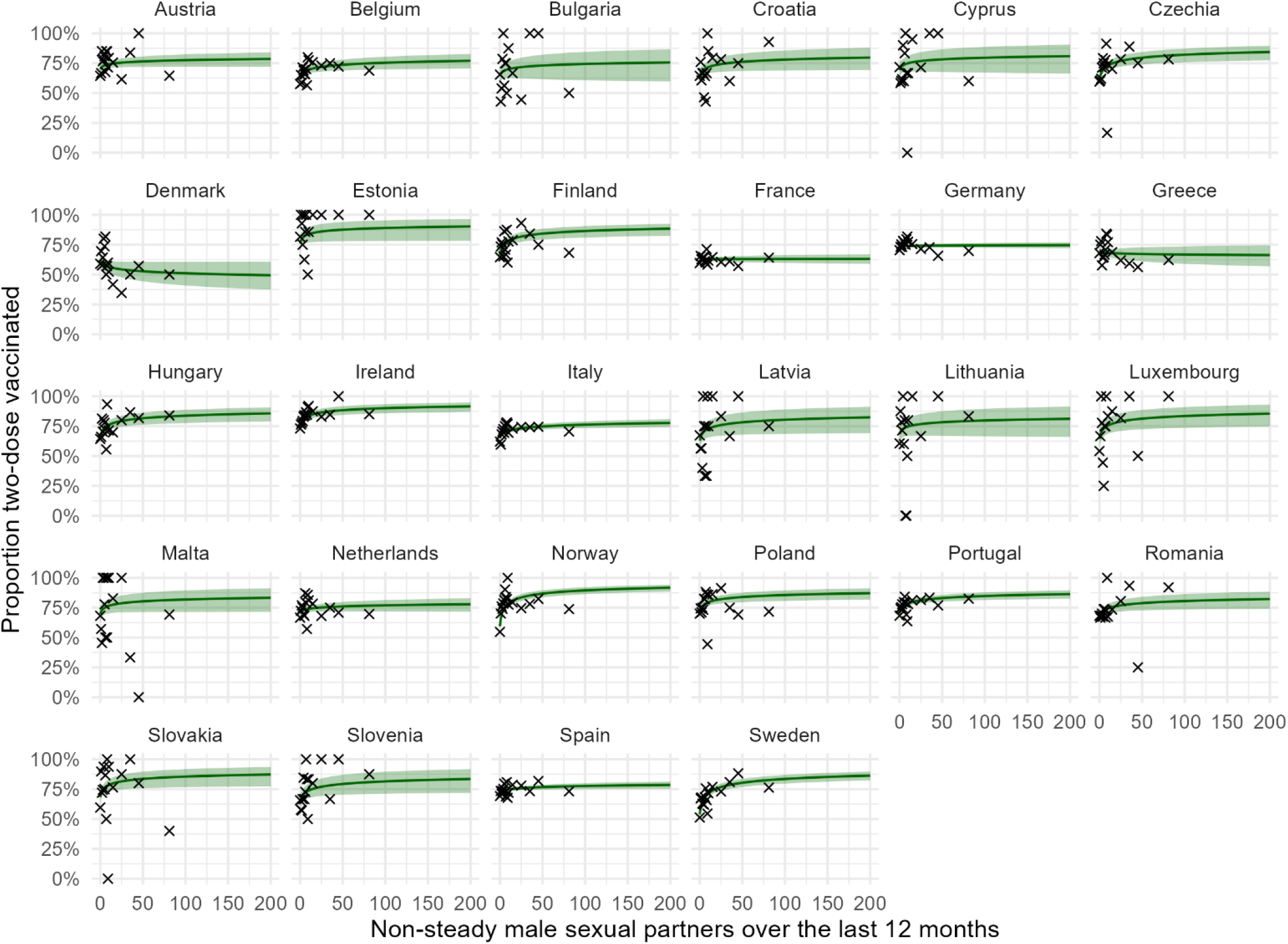
Estimated two-dose vaccine uptake among MSM (increased vaccination scenario A). The probability of being vaccinated against mpox versus the number of non-steady male sexual partners over the last 12 months (or the degree) for the increased vaccination scenario A. The green line and green areas show the median and the 95% uncertainty interval of the posterior vaccination probability of the fitted BME (eq.A1), see Table A3 for more details on the posterior coefficients. The black crosses show the empirical fractions of vaccinated individuals by degree, where banded EMIS-2024 responses were mapped to numeric values using a Weibull degree distribution fitted to the data, providing a model-based alternative to the empirical approach described by Mendez-Lopez et al. [16]. Note that the denominator was small for some data points (which is not visible since the crosses are fractions), and therefore a worse fit to these data points did not influence the posterior as much as data points with a large denominator.

**Figure A5.**
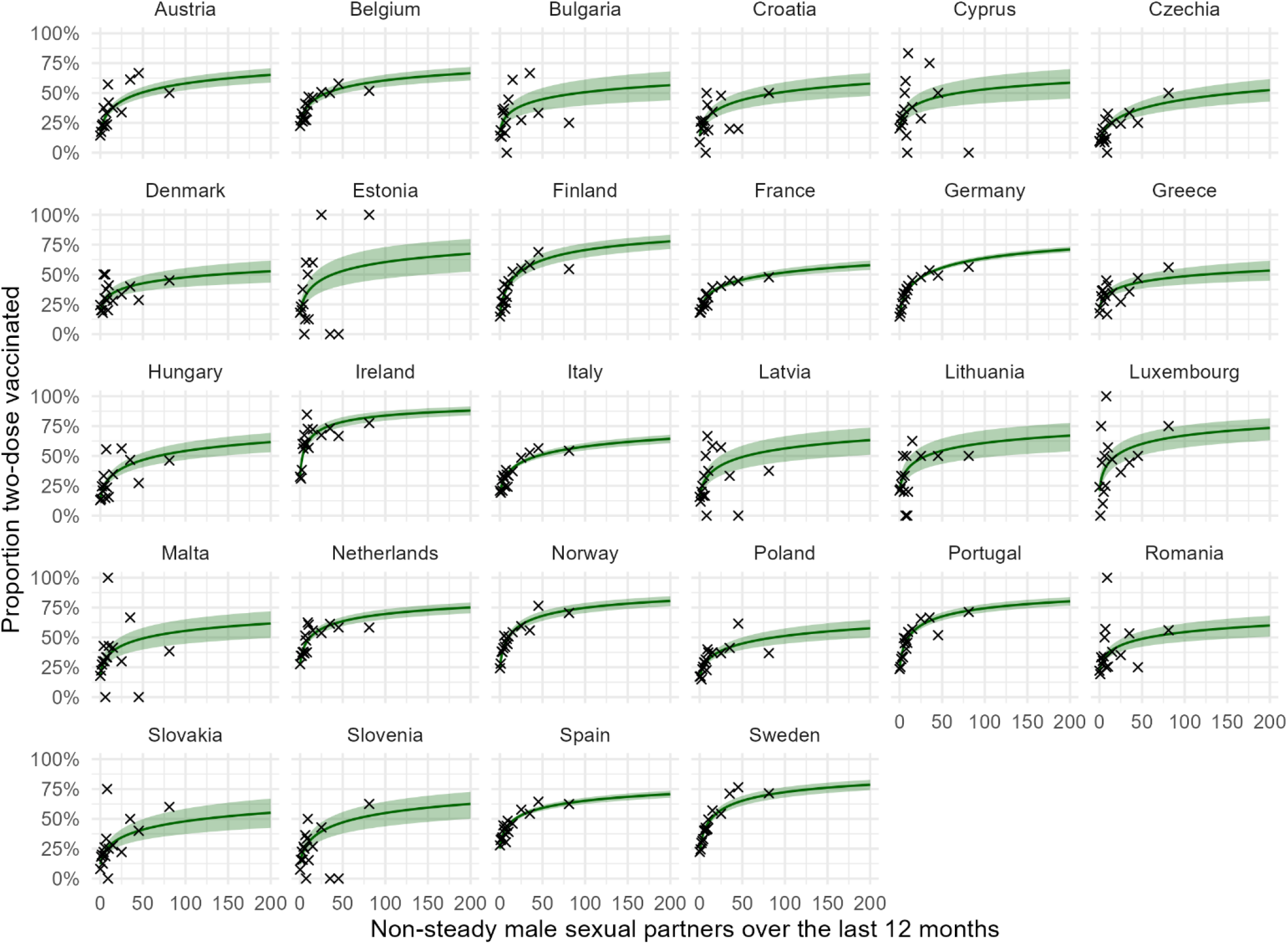
Estimated two-dose vaccine uptake among MSM (increased vaccination scenario B). The probability of being vaccinated against mpox versus the number of non-steady male sexual partners over the last 12 months (or the degree) for the increased vaccination scenario B. The green line and green areas show the median and the 95% uncertainty interval of the posterior vaccination probability of the fitted BME (eq.A1), see Table A4 for more details on the posterior coefficients. The black crosses show the empirical fractions of vaccinated individuals by degree, where banded EMIS-2024 responses were mapped to numeric values using a Weibull degree distribution fitted to the data, providing a model-based alternative to the empirical approach described by Mendez-Lopez et al. [16]. Note that the denominator was small for some data points (which is not visible since the crosses are fractions), and therefore a worse fit to these data points did not influence the posterior as much as data points with a large denominator.

**Table A1.** Model coefficients for estimating the conditional one-dose vaccine uptake among MSM. The country-specific intercept and slope of the fitted BME (eq.A1) for the vaccination probability for the baseline vaccination scenario. Each cell shows the posterior median and the 95% uncertainty interval of the respective parameter.

| Country $c(i)$ | Intercept $\beta_0 + \gamma_{0,c(i)}$ | Slope $\beta_1 + \gamma_{1,c(i)}$ |
| --- | --- | --- |
| Austria | -4.15 (-4.7, -3.66) | 0.67 (0.5, 0.86) |
| Belgium | -3.44 (-3.91, -2.99) | 0.64 (0.49, 0.8) |
| Bulgaria | -5.69 (-7.26, -4.42) | 0.38 (-0.07, 0.72) |
| Croatia | -4.82 (-5.74, -3.94) | 0.48 (0.16, 0.75) |
| Cyprus | -6.19 (-8.13, -4.71) | 0.39 (-0.08, 0.76) |
| Czechia | -5.34 (-6.16, -4.56) | 0.43 (0.08, 0.7) |
| Denmark | -2.45 (-2.93, -2.01) | 0.77 (0.61, 0.96) |
| Estonia | -5.02 (-6.35, -3.91) | 0.48 (0.09, 0.78) |
| Finland | -4.49 (-5.19, -3.92) | 0.83 (0.61, 1.11) |
| France | -2.66 (-2.89, -2.43) | 0.63 (0.55, 0.71) |
| Germany | -3.58 (-3.75, -3.43) | 0.71 (0.66, 0.77) |
| Greece | -5.18 (-6.03, -4.35) | 0.4 (0.09, 0.66) |
| Hungary | -5.93 (-7.05, -4.99) | 0.49 (0.16, 0.8) |
| Ireland | -3.85 (-4.4, -3.29) | 0.49 (0.26, 0.69) |
| Italy | -4.11 (-4.46, -3.77) | 0.6 (0.48, 0.71) |
| Latvia | -4.85 (-6.08, -3.77) | 0.43 (0.02, 0.73) |
| Lithuania | -5.9 (-7.89, -4.4) | 0.43 (-0.03, 0.78) |
| Luxembourg | -4.32 (-5.42, -3.36) | 0.59 (0.3, 0.87) |
| Malta | -4.66 (-5.67, -3.77) | 0.57 (0.29, 0.84) |
| Netherlands | -3.44 (-3.85, -3.06) | 0.74 (0.6, 0.89) |
| Norway | -4.6 (-5.07, -4.19) | 0.7 (0.53, 0.88) |
| Poland | -5.9 (-6.89, -5.08) | 0.58 (0.31, 0.89) |
| Portugal | -4.1 (-4.53, -3.69) | 0.64 (0.49, 0.79) |
| Romania | -4.95 (-5.79, -4.11) | 0.37 (0.01, 0.64) |
| Slovakia | -6.54 (-8.44, -5.14) | 0.37 (-0.12, 0.76) |
| Slovenia | -4.28 (-5.14, -3.48) | 0.57 (0.28, 0.84) |
| Spain | -3.51 (-3.8, -3.21) | 0.52 (0.42, 0.61) |
| Sweden | -3.94 (-4.28, -3.64) | 0.72 (0.59, 0.85) |

**Table A2.** Model coefficients for estimating the two-dose vaccine uptake among MSM (baseline scenario). The country-specific intercept and slope of the fitted BME (eq.A1) for the vaccination probability for the baseline vaccination scenario. Each cell shows the posterior median and the 95% uncertainty interval of the respective parameter.

| Country $c(i)$ | Intercept $\beta_0 + \gamma_{0,c(i)}$ | Slope $\beta_1 + \gamma_{1,c(i)}$ |
| --- | --- | --- |
| Austria | -2.88 (-3.23, -2.54) | 0.62 (0.5, 0.74) |
| Belgium | -2.62 (-2.98, -2.28) | 0.55 (0.43, 0.67) |
| Bulgaria | -5.02 (-6.29, -3.94) | 0.55 (0.21, 0.87) |
| Croatia | -4.01 (-4.74, -3.31) | 0.64 (0.4, 0.88) |
| Cyprus | -4.85 (-6.13, -3.72) | 0.44 (0.06, 0.78) |
| Czechia | -5.23 (-5.96, -4.57) | 0.93 (0.71, 1.18) |
| Denmark | -2.85 (-3.42, -2.33) | 0.44 (0.26, 0.62) |
| Estonia | -5.33 (-6.91, -4.03) | 0.66 (0.28, 1.03) |
| Finland | -3.9 (-4.38, -3.48) | 1.02 (0.86, 1.2) |
| France | -2.2 (-2.39, -2.01) | 0.46 (0.4, 0.52) |
| Germany | -2.79 (-2.9, -2.67) | 0.64 (0.6, 0.69) |
| Greece | -5.29 (-6.33, -4.36) | 0.32 (-0.04, 0.63) |
| Hungary | -5.33 (-6.12, -4.62) | 0.84 (0.61, 1.08) |
| Ireland | -2.11 (-2.4, -1.83) | 0.64 (0.52, 0.76) |
| Italy | -3.16 (-3.39, -2.93) | 0.61 (0.53, 0.68) |
| Latvia | -4.53 (-5.58, -3.6) | 0.76 (0.48, 1.05) |
| Lithuania | -3.2 (-4.08, -2.39) | 0.5 (0.2, 0.78) |
| Luxembourg | -2.75 (-3.55, -1.99) | 0.67 (0.42, 0.92) |
| Malta | -4.77 (-5.92, -3.75) | 0.55 (0.24, 0.84) |
| Netherlands | -2.49 (-2.79, -2.2) | 0.54 (0.43, 0.65) |
| Norway | -3.88 (-4.2, -3.59) | 0.92 (0.8, 1.04) |
| Poland | -5.97 (-6.92, -5.11) | 0.62 (0.34, 0.9) |
| Portugal | -2.52 (-2.75, -2.3) | 0.63 (0.54, 0.72) |
| Romania | -5.49 (-6.49, -4.6) | 0.53 (0.22, 0.82) |
| Slovakia | -6.29 (-7.71, -5.1) | 0.73 (0.37, 1.09) |
| Slovenia | -3.41 (-4.11, -2.75) | 0.77 (0.52, 1.03) |
| Spain | -2.62 (-2.82, -2.42) | 0.48 (0.42, 0.54) |
| Sweden | -3.69 (-3.98, -3.42) | 0.93 (0.83, 1.04) |

**Table A3.**
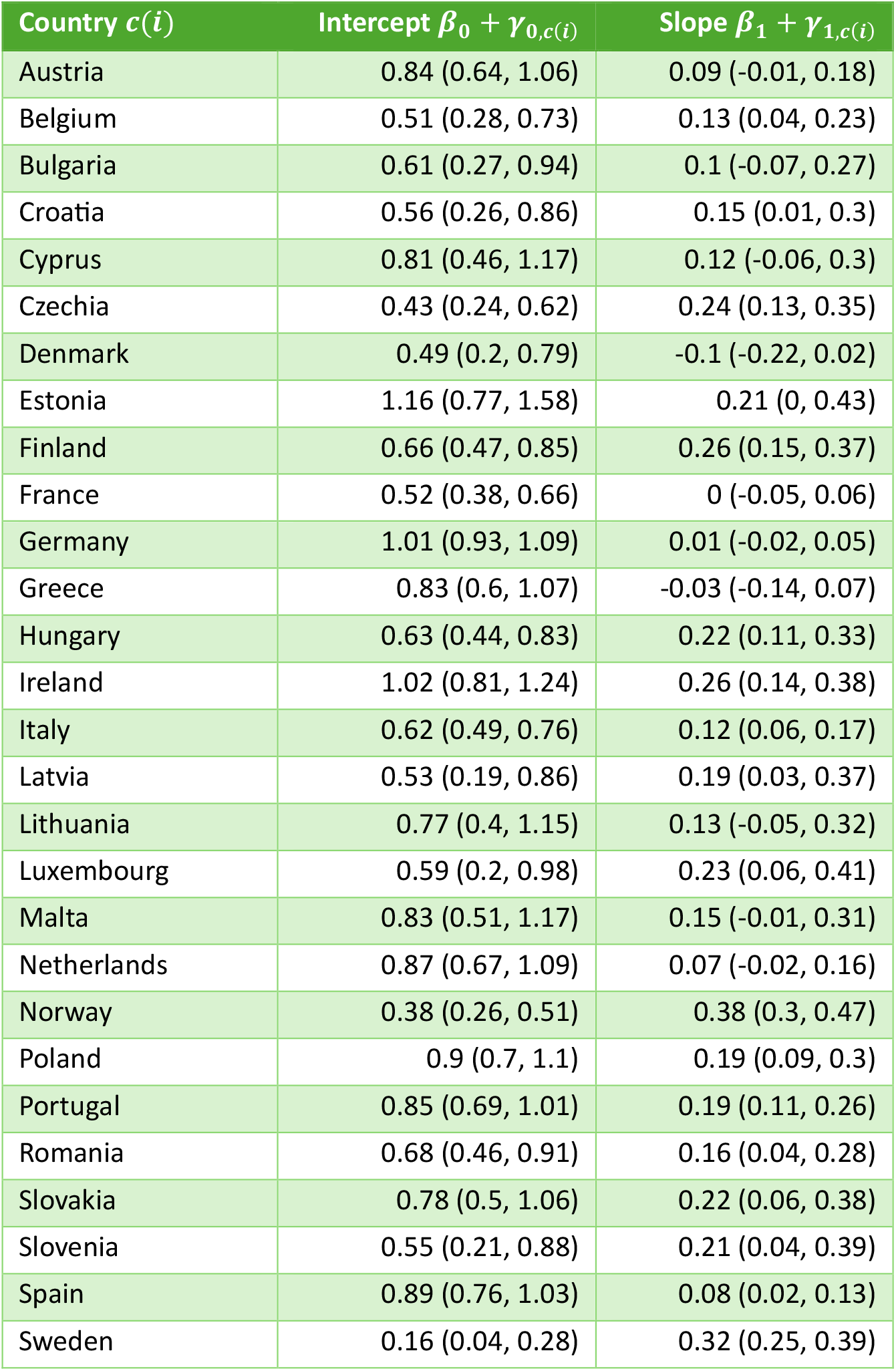
Model coefficients for estimating the two-dose vaccine uptake among MSM (increased vaccination scenario A). The country-specific intercept and slope of the fitted BME (eq.A1) for the vaccination probability for the increased vaccination scenario A. Each cell shows the posterior median and the 95% uncertainty interval of the respective parameter.

| Country $c(i)$ | Intercept $\beta_0 + \gamma_{0,c(i)}$ | Slope $\beta_1 + \gamma_{1,c(i)}$ |
| --- | --- | --- |
| Austria | 0.84 (0.64, 1.06) | 0.09 (-0.01, 0.18) |
| Belgium | 0.51 (0.28, 0.73) | 0.13 (0.04, 0.23) |
| Bulgaria | 0.61 (0.27, 0.94) | 0.1 (-0.07, 0.27) |
| Croatia | 0.56 (0.26, 0.86) | 0.15 (0.01, 0.3) |
| Cyprus | 0.81 (0.46, 1.17) | 0.12 (-0.06, 0.3) |
| Czechia | 0.43 (0.24, 0.62) | 0.24 (0.13, 0.35) |
| Denmark | 0.49 (0.2, 0.79) | -0.1 (-0.22, 0.02) |
| Estonia | 1.16 (0.77, 1.58) | 0.21 (0, 0.43) |
| Finland | 0.66 (0.47, 0.85) | 0.26 (0.15, 0.37) |
| France | 0.52 (0.38, 0.66) | 0 (-0.05, 0.06) |
| Germany | 1.01 (0.93, 1.09) | 0.01 (-0.02, 0.05) |
| Greece | 0.83 (0.6, 1.07) | -0.03 (-0.14, 0.07) |
| Hungary | 0.63 (0.44, 0.83) | 0.22 (0.11, 0.33) |
| Ireland | 1.02 (0.81, 1.24) | 0.26 (0.14, 0.38) |
| Italy | 0.62 (0.49, 0.76) | 0.12 (0.06, 0.17) |
| Latvia | 0.53 (0.19, 0.86) | 0.19 (0.03, 0.37) |
| Lithuania | 0.77 (0.4, 1.15) | 0.13 (-0.05, 0.32) |
| Luxembourg | 0.59 (0.2, 0.98) | 0.23 (0.06, 0.41) |
| Malta | 0.83 (0.51, 1.17) | 0.15 (-0.01, 0.31) |
| Netherlands | 0.87 (0.67, 1.09) | 0.07 (-0.02, 0.16) |
| Norway | 0.38 (0.26, 0.51) | 0.38 (0.3, 0.47) |
| Poland | 0.9 (0.7, 1.1) | 0.19 (0.09, 0.3) |
| Portugal | 0.85 (0.69, 1.01) | 0.19 (0.11, 0.26) |
| Romania | 0.68 (0.46, 0.91) | 0.16 (0.04, 0.28) |
| Slovakia | 0.78 (0.5, 1.06) | 0.22 (0.06, 0.38) |
| Slovenia | 0.55 (0.21, 0.88) | 0.21 (0.04, 0.39) |
| Spain | 0.89 (0.76, 1.03) | 0.08 (0.02, 0.13) |
| Sweden | 0.16 (0.04, 0.28) | 0.32 (0.25, 0.39) |

**Table A4.** Model coefficients for estimating the two-dose vaccine uptake among MSM (increased vaccination scenario B). The country-specific intercept and slope of the fitted BME (eq.A1) for the vaccination probability for the increased vaccination scenario B. Each cell shows the posterior median and the 95% uncertainty interval of the respective parameter.

| Country $c(i)$ | Intercept $\beta_0 + \gamma_{0,c(i)}$ | Slope $\beta_1 + \gamma_{1,c(i)}$ |
| --- | --- | --- |
| Austria | -1.71 (-1.91, -1.51) | 0.44 (0.37, 0.51) |
| Belgium | -1.31 (-1.51, -1.1) | 0.38 (0.3, 0.44) |
| Bulgaria | -1.57 (-1.92, -1.22) | 0.35 (0.23, 0.46) |
| Croatia | -1.81 (-2.12, -1.51) | 0.4 (0.3, 0.5) |
| Cyprus | -1.38 (-1.73, -1.02) | 0.32 (0.2, 0.44) |
| Czechia | -2.34 (-2.59, -2.1) | 0.46 (0.36, 0.56) |
| Denmark | -1.46 (-1.76, -1.16) | 0.3 (0.19, 0.39) |
| Estonia | -1.63 (-2.04, -1.25) | 0.45 (0.31, 0.59) |
| Finland | -1.69 (-1.89, -1.49) | 0.55 (0.47, 0.64) |
| France | -1.58 (-1.72, -1.43) | 0.36 (0.31, 0.41) |
| Germany | -1.69 (-1.78, -1.61) | 0.49 (0.46, 0.52) |
| Greece | -1.33 (-1.56, -1.11) | 0.28 (0.19, 0.37) |
| Hungary | -1.87 (-2.09, -1.66) | 0.44 (0.36, 0.53) |
| Ireland | -0.7 (-0.88, -0.52) | 0.51 (0.43, 0.59) |
| Italy | -1.51 (-1.64, -1.38) | 0.4 (0.35, 0.44) |
| Latvia | -1.7 (-2.06, -1.34) | 0.42 (0.31, 0.53) |
| Lithuania | -1.35 (-1.75, -0.96) | 0.39 (0.27, 0.5) |
| Luxembourg | -1.33 (-1.73, -0.93) | 0.44 (0.33, 0.55) |
| Malta | -1.49 (-1.84, -1.14) | 0.37 (0.26, 0.48) |
| Netherlands | -1.01 (-1.19, -0.83) | 0.4 (0.33, 0.46) |
| Norway | -1.15 (-1.27, -1.03) | 0.49 (0.42, 0.55) |
| Poland | -1.68 (-1.88, -1.48) | 0.37 (0.29, 0.45) |
| Portugal | -1.18 (-1.33, -1.04) | 0.49 (0.43, 0.55) |
| Romania | -1.34 (-1.56, -1.11) | 0.33 (0.23, 0.42) |
| Slovakia | -2 (-2.32, -1.7) | 0.42 (0.3, 0.54) |
| Slovenia | -1.92 (-2.29, -1.59) | 0.46 (0.34, 0.57) |
| Spain | -1.1 (-1.22, -0.97) | 0.37 (0.33, 0.42) |
| Sweden | -1.31 (-1.43, -1.18) | 0.49 (0.43, 0.55) |

**Figure A6.**
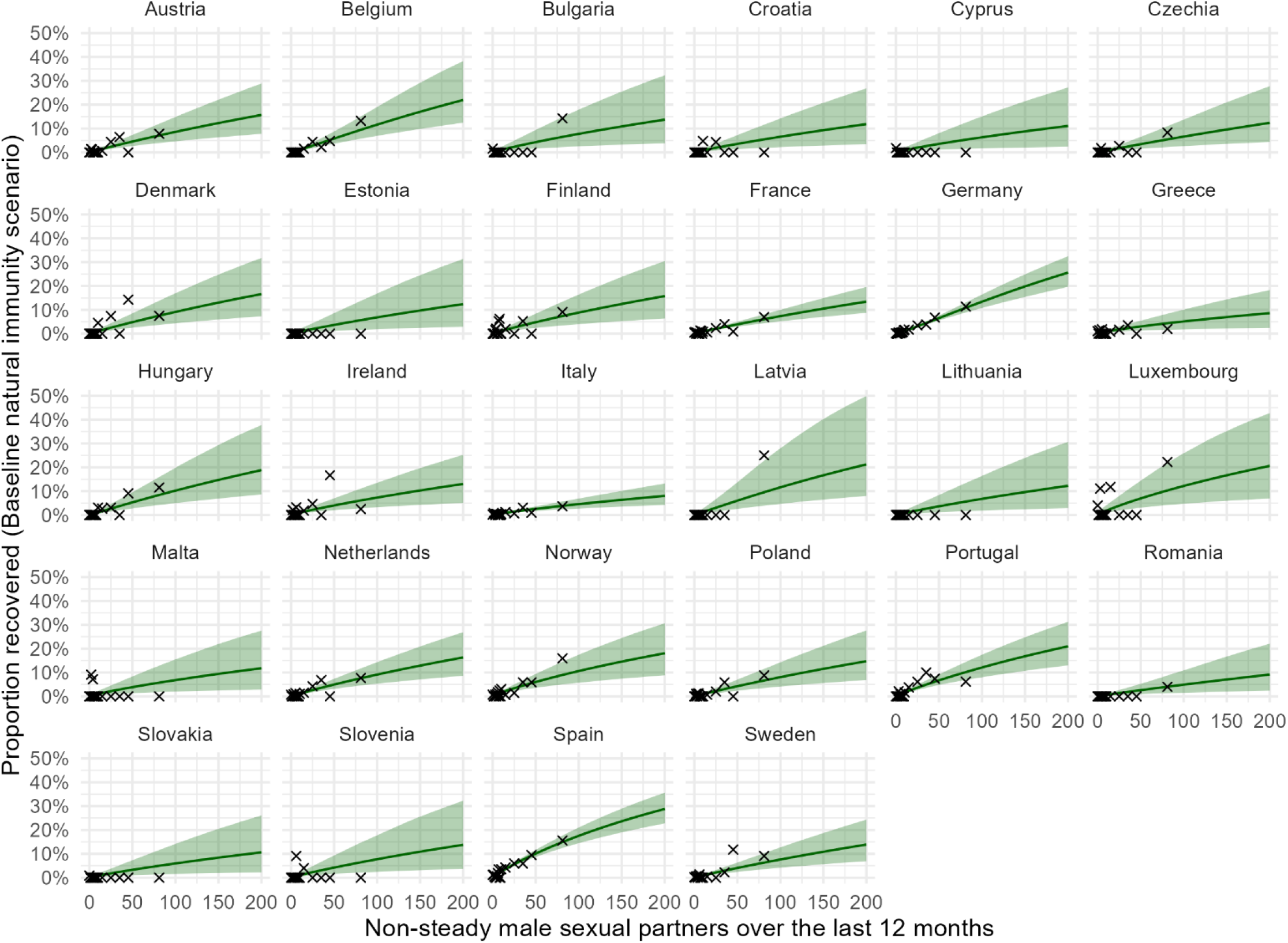
Estimated proportion of MSM recovered from mpox (baseline natural immunity scenario). The probability of being recovered from mpox versus the number of non-steady male sexual partners over the last 12 months (or the degree) for the baseline natural immunity scenario. The green line and green areas show the median and the 95% uncertainty interval of the posterior vaccination probability of the fitted BME (eq.A1), see Table A5 for more details on the posterior coefficients. The black crosses show the empirical fractions of vaccinated individuals by degree, where banded EMIS-2024 responses were mapped to numeric values using a Weibull degree distribution fitted to the data, providing a model-based alternative to the empirical approach described by Mendez-Lopez et al. [16]. Note that the denominator was small for some data points (which is not visible since the crosses are fractions), and therefore a worse fit to these data points did not influence the posterior as much as data points with a large denominator.

**Table A5.** Model coefficients for estimating the proportion of MSM recovered from mpox (baseline natural immunity scenario). The country-specific intercept and slope of the fitted BME (eq.A1) for the probability of being recovered for the baseline natural immunity scenario. Each cell shows the posterior median and the 95% uncertainty interval of the respective parameter.

| Country $c(i)$ | Intercept $\beta_0 + \gamma_{0,c(i)}$ | Slope $\beta_1 + \gamma_{1,c(i)}$ |
| --- | --- | --- |
| Bulgaria | -6.70 (-7.92, -5.43) | 0.91 (0.52, 1.24) |
| Croatia | -6.99 (-8.28, -5.81) | 0.93 (0.55, 1.27) |
| Cyprus | -6.73 (-7.99, -5.35) | 0.86 (0.41, 1.19) |
| Czechia | -7.30 (-8.60, -6.29) | 1.00 (0.68, 1.37) |
| Denmark | -6.89 (-8.12, -5.81) | 0.99 (0.69, 1.32) |
| Estonia | -7.00 (-8.43, -5.75) | 0.94 (0.55, 1.31) |
| Finland | -6.72 (-7.71, -5.77) | 0.95 (0.64, 1.23) |
| France | -7.03 (-7.83, -6.30) | 0.97 (0.77, 1.19) |
| Germany | -7.22 (-7.82, -6.65) | 1.16 (1.00, 1.32) |
| Greece | -6.54 (-7.59, -5.29) | 0.77 (0.35, 1.07) |
| Hungary | -7.00 (-8.22, -6.05) | 1.05 (0.77, 1.39) |
| Ireland | -6.80 (-7.82, -5.77) | 0.92 (0.60, 1.20) |
| Italy | -7.04 (-7.91, -6.18) | 0.87 (0.60, 1.10) |
| Latvia | -6.90 (-8.37, -5.74) | 1.06 (0.74, 1.48) |
| Lithuania | -7.00 (-8.44, -5.71) | 0.94 (0.54, 1.31) |
| Luxembourg | -6.11 (-7.26, -4.59) | 0.89 (0.46, 1.21) |
| Malta | -6.60 (-7.80, -5.20) | 0.85 (0.41, 1.17) |
| Netherlands | -6.73 (-7.63, -5.85) | 0.96 (0.70, 1.21) |
| Norway | -6.27 (-6.98, -5.51) | 0.89 (0.63, 1.12) |
| Poland | -7.03 (-8.10, -6.12) | 0.99 (0.72, 1.29) |
| Portugal | -6.37 (-7.11, -5.61) | 0.95 (0.73, 1.16) |
| Romania | -7.44 (-8.93, -6.28) | 0.96 (0.58, 1.35) |
| Slovakia | -6.91 (-8.17, -5.71) | 0.89 (0.46, 1.22) |
| Slovenia | -6.78 (-8.01, -5.54) | 0.93 (0.54, 1.26) |
| Spain | -5.87 (-6.44, -5.31) | 0.94 (0.78, 1.09) |
| Sweden | -6.99 (-7.87, -6.22) | 0.97 (0.73, 1.23) |
| Bulgaria | -6.70 (-7.92, -5.43) | 0.91 (0.52, 1.24) |
| Croatia | -6.99 (-8.28, -5.81) | 0.93 (0.55, 1.27) |

**Figure A7.**
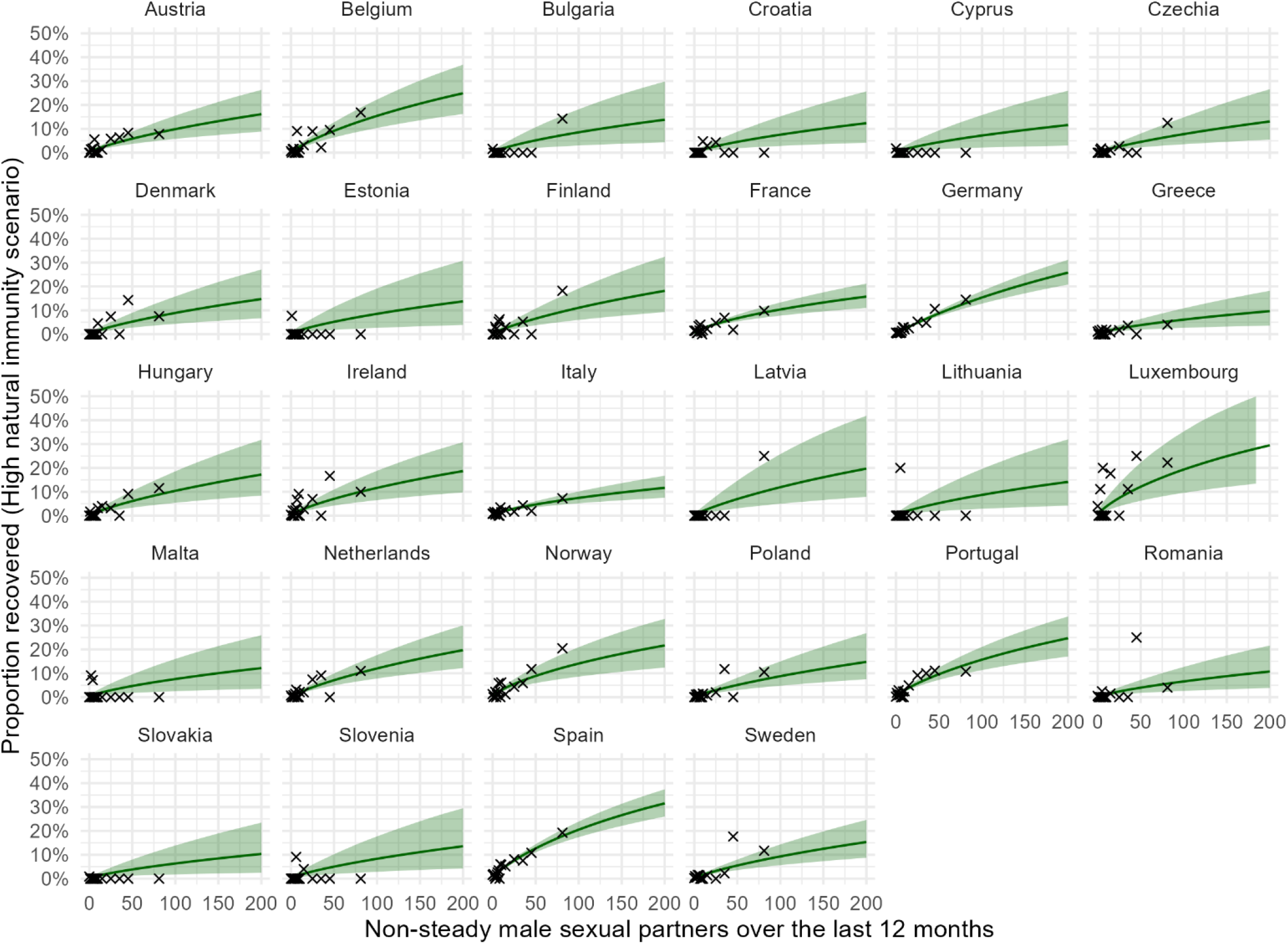
Estimated proportion of MSM recovered from mpox (high natural immunity scenario). The probability of being recovered from mpox versus the number of non-steady male sexual partners over the last 12 months (or the degree) for the high natural immunity scenario. The green line and green areas show the median and the 95% uncertainty interval of the posterior vaccination probability of the fitted BME (eq.A1), see Table A6 for more details on the posterior coefficients. The black crosses show the empirical fractions of vaccinated individuals by degree, where banded EMIS-2024 responses were mapped to numeric values using a Weibull degree distribution fitted to the data, providing a model-based alternative to the empirical approach described by Mendez-Lopez et al. [16]. Note that the denominator was small for some data points (which is not visible since the crosses are fractions), and therefore a worse fit to these data points did not influence the posterior as much as data points with a large denominator.

**Table A6.**
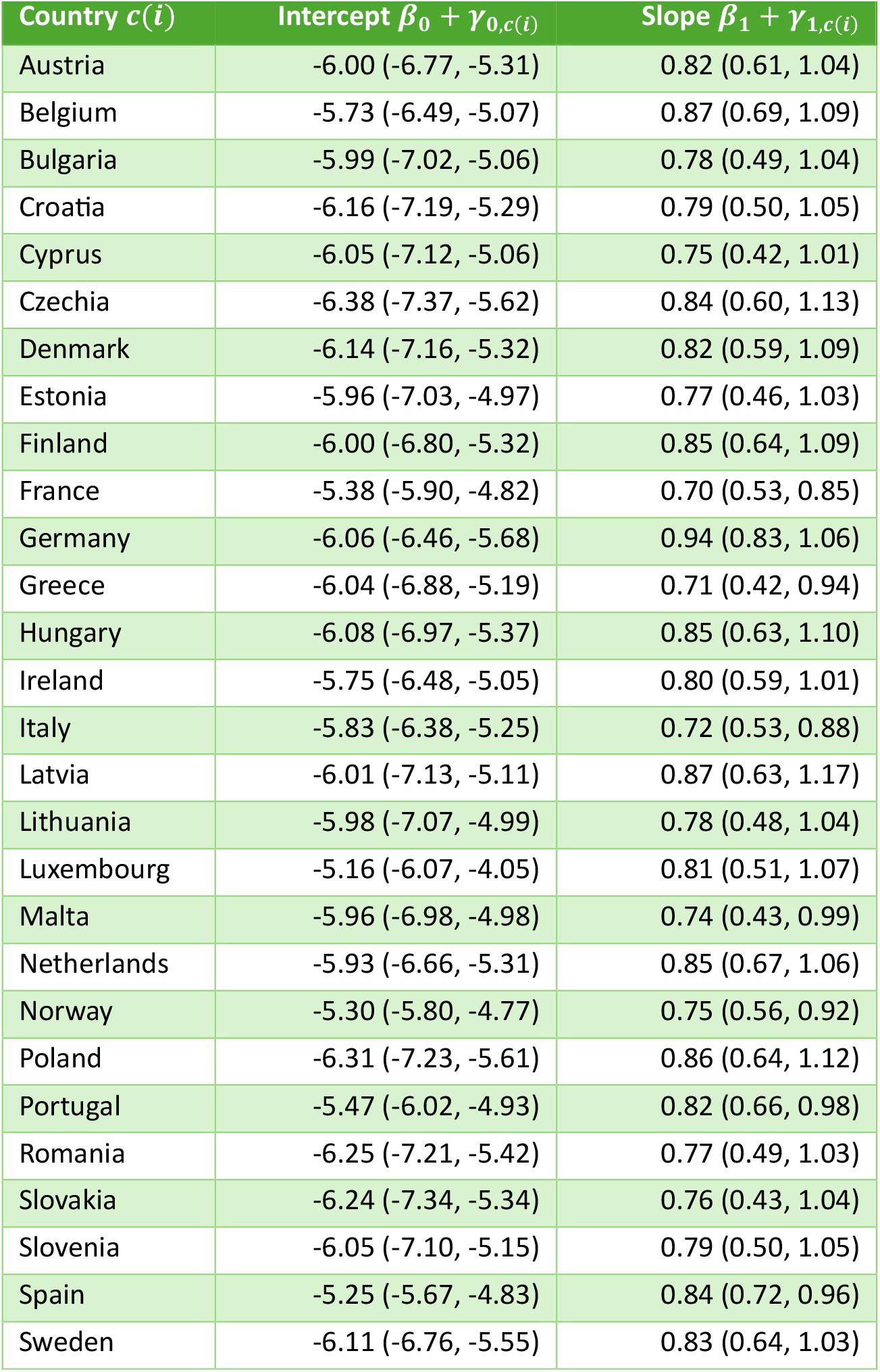
Model coefficients for estimating the proportion of MSM recovered from mpox (high natural immunity scenario). The country-specific intercept and slope of the fitted BME (eq.A1) for the probability of being recovered for the high natural immunity scenario. Each cell shows the posterior median and the 95% uncertainty interval of the respective parameter.

| Country $c(i)$ | Intercept $\beta_0 + \gamma_{0,c(i)}$ | Slope $\beta_1 + \gamma_{1,c(i)}$ |
| --- | --- | --- |
| Austria | -6.00 (-6.77, -5.31) | 0.82 (0.61, 1.04) |
| Belgium | -5.73 (-6.49, -5.07) | 0.87 (0.69, 1.09) |
| Bulgaria | -5.99 (-7.02, -5.06) | 0.78 (0.49, 1.04) |
| Croatia | -6.16 (-7.19, -5.29) | 0.79 (0.50, 1.05) |
| Cyprus | -6.05 (-7.12, -5.06) | 0.75 (0.42, 1.01) |
| Czechia | -6.38 (-7.37, -5.62) | 0.84 (0.60, 1.13) |
| Denmark | -6.14 (-7.16, -5.32) | 0.82 (0.59, 1.09) |
| Estonia | -5.96 (-7.03, -4.97) | 0.77 (0.46, 1.03) |
| Finland | -6.00 (-6.80, -5.32) | 0.85 (0.64, 1.09) |
| France | -5.38 (-5.90, -4.82) | 0.70 (0.53, 0.85) |
| Germany | -6.06 (-6.46, -5.68) | 0.94 (0.83, 1.06) |
| Greece | -6.04 (-6.88, -5.19) | 0.71 (0.42, 0.94) |
| Hungary | -6.08 (-6.97, -5.37) | 0.85 (0.63, 1.10) |
| Ireland | -5.75 (-6.48, -5.05) | 0.80 (0.59, 1.01) |
| Italy | -5.83 (-6.38, -5.25) | 0.72 (0.53, 0.88) |
| Latvia | -6.01 (-7.13, -5.11) | 0.87 (0.63, 1.17) |
| Lithuania | -5.98 (-7.07, -4.99) | 0.78 (0.48, 1.04) |
| Luxembourg | -5.16 (-6.07, -4.05) | 0.81 (0.51, 1.07) |
| Malta | -5.96 (-6.98, -4.98) | 0.74 (0.43, 0.99) |
| Netherlands | -5.93 (-6.66, -5.31) | 0.85 (0.67, 1.06) |
| Norway | -5.30 (-5.80, -4.77) | 0.75 (0.56, 0.92) |
| Poland | -6.31 (-7.23, -5.61) | 0.86 (0.64, 1.12) |
| Portugal | -5.47 (-6.02, -4.93) | 0.82 (0.66, 0.98) |
| Romania | -6.25 (-7.21, -5.42) | 0.77 (0.49, 1.03) |
| Slovakia | -6.24 (-7.34, -5.34) | 0.76 (0.43, 1.04) |
| Slovenia | -6.05 (-7.10, -5.15) | 0.79 (0.50, 1.05) |
| Spain | -5.25 (-5.67, -4.83) | 0.84 (0.72, 0.96) |
| Sweden | -6.11 (-6.76, -5.55) | 0.83 (0.64, 1.03) |

**Table A7.** Estimated two-dose vaccine uptake among MSM. The vaccine uptake aggregated over all MSM (any degree) for each of the three vaccination scenarios (Table 1), resulting from the fitted Weibull degree distribution and the respective BEM. Each cell shows the posterior median and the 95% uncertainty interval of the vaccine uptake among MSM. This table corresponds to assuming a maximum number of 200 non-steady partners per year, please see Table A9 for a sensitivity analysis considering 350 non-steady partners per year.

| Country | Baseline vaccination scenario | Increased vaccination scenario A | Increased vaccination scenario B |
| --- | --- | --- | --- |
| Austria | 18.00% (15.18%, 21.09%) | 75.00% (71.35%, 78.47%) | 30.88% (27.73%, 34.16%) |
| Belgium | 19.26% (16.27%, 22.59%) | 69.57% (65.42%, 73.46%) | 36.60% (33.13%, 40.20%) |
| Bulgaria | 2.41% (0.78%, 5.75%) | 70.54% (62.47%, 77.59%) | 29.76% (23.53%, 36.60%) |
| Croatia | 7.50% (4.58%, 11.44%) | 71.53% (65.38%, 77.21%) | 27.33% (22.45%, 32.40%) |
| Cyprus | 2.17% (0.64%, 5.49%) | 75.50% (67.78%, 82.30%) | 32.83% (26.14%, 40.19%) |
| Czechia | 5.42% (3.57%, 7.81%) | 71.78% (67.61%, 75.65%) | 20.43% (17.30%, 23.76%) |
| Denmark | 13.01% (9.37%, 17.38%) | 59.32% (53.06%, 65.34%) | 29.83% (25.08%, 35.03%) |
| Estonia | 2.41% (0.55%, 6.96%) | 84.00% (76.43%, 90.25%) | 32.67% (24.96%, 41.05%) |
| Finland | 18.35% (15.11%, 21.81%) | 77.03% (73.11%, 80.56%) | 36.24% (32.55%, 40.03%) |
| France | 22.63% (20.83%, 24.52%) | 64.49% (62.12%, 66.82%) | 30.05% (28.09%, 32.07%) |
| Germany | 20.21% (19.22%, 21.24%) | 75.72% (74.47%, 76.93%) | 33.24% (32.05%, 34.44%) |
| Greece | 1.01% (0.44%, 2.03%) | 70.38% (65.96%, 74.49%) | 31.68% (27.91%, 35.67%) |
| Hungary | 3.82% (2.31%, 5.91%) | 75.29% (71.08%, 79.16%) | 27.77% (24.14%, 31.59%) |
| Ireland | 31.38% (27.44%, 35.46%) | 83.18% (79.68%, 86.40%) | 56.44% (52.56%, 60.36%) |
| Italy | 14.23% (12.85%, 15.73%) | 71.56% (69.32%, 73.72%) | 33.06% (31.16%, 35.02%) |
| Latvia | 6.36% (2.97%, 11.79%) | 72.18% (64.28%, 79.18%) | 30.42% (23.86%, 37.39%) |
| Lithuania | 11.02% (5.38%, 19.25%) | 75.08% (66.52%, 82.70%) | 36.21% (28.21%, 44.60%) |
| Luxembourg | 21.77% (14.59%, 30.51%) | 74.64% (66.58%, 81.73%) | 39.12% (31.48%, 47.18%) |
| Malta | 3.02% (1.14%, 6.46%) | 76.78% (69.85%, 82.95%) | 32.43% (26.00%, 39.29%) |
| Netherlands | 20.80% (18.07%, 23.74%) | 75.08% (71.69%, 78.30%) | 44.43% (41.21%, 47.70%) |
| Norway | 15.23% (12.99%, 17.60%) | 75.37% (72.64%, 77.97%) | 45.05% (42.25%, 47.91%) |
| Poland | 1.14% (0.56%, 2.05%) | 79.45% (76.03%, 82.60%) | 28.75% (25.60%, 32.04%) |
| Portugal | 23.73% (21.37%, 26.22%) | 78.45% (75.80%, 80.94%) | 44.46% (41.81%, 47.16%) |
| Romania | 1.43% (0.62%, 2.82%) | 74.33% (69.74%, 78.53%) | 33.85% (29.63%, 38.21%) |
| Slovakia | 1.16% (0.30%, 3.17%) | 77.98% (72.18%, 83.11%) | 24.41% (19.54%, 29.60%) |
| Slovenia | 16.13% (10.70%, 22.65%) | 73.05% (65.85%, 79.58%) | 27.50% (21.75%, 33.51%) |
| Spain | 17.04% (15.57%, 18.61%) | 75.67% (73.63%, 77.65%) | 41.32% (39.31%, 43.35%) |
| Sweden | 18.02% (15.90%, 20.27%) | 68.77% (65.98%, 71.47%) | 41.78% (39.15%, 44.47%) |

**Table A8.** Estimated proportion of recovered MSM. The proportion of MSM recovered from mpox aggregated over all degrees for each of the two natural immunity scenarios (Table 2), resulting from the fitted Weibull degree distribution and the respective BEM. Each cell shows the posterior median and the 95% uncertainty interval of the proportion of recovered MSM. This table corresponds to assuming a maximum number of 200 non-steady partners per year, please see Table A10 for a sensitivity analysis considering 350 non-steady partners per year.

| Country | Baseline natural immunity scenario | High natural immunity scenario |
| --- | --- | --- |
| Austria | 1.33% (0.75%, 2.19%) | 1.94% (1.20%, 2.95%) |
| Belgium | 1.58% (0.93%, 2.50%) | 2.91% (1.97%, 4.16%) |
| Bulgaria | 1.31% (0.51%, 2.91%) | 1.75% (0.71%, 3.58%) |
| Croatia | 1.07% (0.42%, 2.21%) | 1.53% (0.66%, 2.93%) |
| Cyprus | 1.12% (0.39%, 2.59%) | 1.52% (0.56%, 3.23%) |
| Czechia | 0.99% (0.44%, 1.89%) | 1.46% (0.73%, 2.55%) |
| Denmark | 1.41% (0.71%, 2.59%) | 1.72% (0.88%, 2.98%) |
| Estonia | 1.10% (0.36%, 2.58%) | 1.77% (0.65%, 3.82%) |
| Finland | 1.44% (0.74%, 2.52%) | 2.11% (1.21%, 3.42%) |
| France | 1.15% (0.82%, 1.56%) | 2.45% (1.91%, 3.14%) |
| Germany | 1.75% (1.43%, 2.12%) | 2.63% (2.24%, 3.06%) |
| Greece | 1.02% (0.48%, 1.91%) | 1.36% (0.71%, 2.29%) |
| Hungary | 1.50% (0.77%, 2.72%) | 1.96% (1.07%, 3.29%) |
| Ireland | 1.22% (0.62%, 2.17%) | 2.35% (1.41%, 3.67%) |
| Italy | 0.81% (0.53%, 1.19%) | 1.68% (1.25%, 2.23%) |
| Latvia | 1.70% (0.71%, 3.86%) | 2.22% (0.98%, 4.58%) |
| Lithuania | 1.09% (0.36%, 2.55%) | 1.78% (0.68%, 3.87%) |
| Luxembourg | 2.15% (0.96%, 4.83%) | 4.08% (2.10%, 7.91%) |
| Malta | 1.21% (0.46%, 2.74%) | 1.63% (0.65%, 3.34%) |
| Netherlands | 1.46% (0.90%, 2.25%) | 2.28% (1.54%, 3.24%) |
| Norway | 1.87% (1.15%, 2.90%) | 3.11% (2.13%, 4.37%) |
| Poland | 1.23% (0.67%, 2.07%) | 1.62% (0.93%, 2.58%) |
| Portugal | 2.01% (1.38%, 2.85%) | 3.19% (2.37%, 4.22%) |
| Romania | 0.76% (0.28%, 1.58%) | 1.35% (0.62%, 2.42%) |
| Slovakia | 1.01% (0.35%, 2.25%) | 1.31% (0.47%, 2.68%) |
| Slovenia | 1.27% (0.49%, 2.82%) | 1.69% (0.67%, 3.39%) |
| Spain | 3.07% (2.50%, 3.75%) | 4.17% (3.51%, 4.93%) |
| Sweden | 1.19% (0.71%, 1.88%) | 1.79% (1.17%, 2.59%) |

## Appendix B. Sensitivity Analysis: Impact of non-steady partner truncation on estimated vaccine uptake and proportion of recovered

In the main analysis, we consider a maximum number of 200 non-steady partners per 12 months, which led to truncating the discrete Weibull degree distribution at 200. However, the maximum number of non-steady partners is an uncertain and possibly impactful parameter. As illustrated e.g. by Mendez-Lopez et al. [16], some MSM may have more than 200 non-steady partners. Hence, we conduct a sensitivity analysis by rerunning the model with *d*_max,2_ = 350 in the following. Tables A9 and A10 below show the resulting estimates for the total vaccine uptake and natural immunity among MSM. Comparing these results to Tables A7 and A8, respectively, which used *d*_max,2_ = 200, we see that there the absolute difference of the median estimates is very small for all countries and all vaccine uptake estimates (less than 0.1 percentage points) and natural immunity estimates (less than 0.03 percentage points). Hence, these results seem robust to changes in the parameter *d*_max,2_. Furthermore, Figure A8 shows that the results on the expected reduction of the reproduction number (Figure 4 in the main text) are not substantially affected by whether the number of non-steady partners is truncated at 200 or 350.

**Table A9.** Estimated two-dose vaccine uptake among MSM. The vaccine uptake aggregated over all MSM (any degree) for each of the three vaccination scenarios (Table 1), resulting from the fitted Weibull degree distribution and the respective BEM. Each cell shows the posterior median and the 95% uncertainty interval of the vaccine uptake among MSM. This table corresponds to assuming a maximum number of 350 non-steady partners per year, as a sensitivity analysis to Table A7 that assumed 200 non-steady partners per year and was used for the main text.

| Country | Baseline vaccination scenario | Increased vaccination scenario A | Increased vaccination scenario B |
| --- | --- | --- | --- |
| Austria | 18.01% (15.20%, 21.10%) | 74.98% (71.34%, 78.45%) | 30.89% (27.72%, 34.12%) |
| Belgium | 19.23% (16.24%, 22.56%) | 69.54% (65.45%, 73.44%) | 36.60% (33.12%, 40.25%) |
| Bulgaria | 2.41% (0.77%, 5.69%) | 70.53% (62.34%, 77.62%) | 29.75% (23.45%, 36.67%) |
| Croatia | 7.51% (4.58%, 11.44%) | 71.53% (65.40%, 77.17%) | 27.28% (22.48%, 32.46%) |
| Cyprus | 2.17% (0.63%, 5.44%) | 75.50% (67.71%, 82.32%) | 32.84% (26.03%, 40.17%) |
| Czechia | 5.42% (3.56%, 7.81%) | 71.78% (67.55%, 75.67%) | 20.42% (17.28%, 23.79%) |
| Denmark | 13.04% (9.42%, 17.47%) | 59.36% (53.08%, 65.38%) | 29.84% (25.02%, 35.05%) |
| Estonia | 2.40% (0.56%, 6.93%) | 83.99% (76.52%, 90.29%) | 32.71% (24.94%, 41.03%) |
| Finland | 18.31% (15.08%, 21.85%) | 77.04% (73.17%, 80.60%) | 36.20% (32.54%, 40.01%) |
| France | 22.63% (20.83%, 24.51%) | 64.50% (62.11%, 66.84%) | 30.06% (28.12%, 32.06%) |
| Germany | 20.20% (19.21%, 21.22%) | 75.72% (74.47%, 76.93%) | 33.23% (32.04%, 34.43%) |
| Greece | 1.01% (0.43%, 2.03%) | 70.38% (65.93%, 74.55%) | 31.69% (27.92%, 35.65%) |
| Hungary | 3.82% (2.29%, 5.89%) | 75.29% (71.13%, 79.20%) | 27.75% (24.12%, 31.56%) |
| Ireland | 31.32% (27.37%, 35.53%) | 83.18% (79.69%, 86.37%) | 56.43% (52.49%, 60.40%) |
| Italy | 14.24% (12.82%, 15.72%) | 71.55% (69.30%, 73.72%) | 33.08% (31.16%, 35.03%) |
| Latvia | 6.37% (2.94%, 11.67%) | 72.17% (64.22%, 79.01%) | 30.41% (23.91%, 37.33%) |
| Lithuania | 11.02% (5.36%, 19.34%) | 75.15% (66.38%, 82.65%) | 36.24% (28.29%, 44.75%) |
| Luxembourg | 21.77% (14.66%, 30.55%) | 74.65% (66.43%, 81.67%) | 39.10% (31.51%, 47.15%) |
| Malta | 3.00% (1.12%, 6.46%) | 76.72% (69.73%, 82.95%) | 32.38% (26.08%, 39.18%) |
| Netherlands | 20.79% (18.07%, 23.76%) | 75.09% (71.68%, 78.35%) | 44.41% (41.21%, 47.73%) |
| Norway | 15.19% (12.98%, 17.60%) | 75.37% (72.61%, 77.96%) | 45.05% (42.25%, 47.89%) |
| Poland | 1.15% (0.57%, 2.07%) | 79.45% (76.04%, 82.61%) | 28.73% (25.57%, 32.01%) |
| Portugal | 23.73% (21.37%, 26.22%) | 78.47% (75.84%, 80.94%) | 44.47% (41.79%, 47.17%) |
| Romania | 1.43% (0.62%, 2.83%) | 74.31% (69.80%, 78.50%) | 33.84% (29.66%, 38.22%) |
| Slovakia | 1.17% (0.31%, 3.18%) | 77.94% (72.12%, 83.11%) | 24.35% (19.47%, 29.59%) |
| Slovenia | 16.04% (10.64%, 22.66%) | 73.12% (65.78%, 79.60%) | 27.46% (21.71%, 33.57%) |
| Spain | 17.03% (15.58%, 18.58%) | 75.69% (73.65%, 77.69%) | 41.31% (39.32%, 43.38%) |
| Sweden | 17.98% (15.87%, 20.22%) | 68.78% (65.96%, 71.44%) | 41.79% (39.15%, 44.44%) |

**Table A10.** Estimated proportion of recovered MSM. The proportion of MSM recovered from mpox aggregated over all degrees for each of the two natural immunity scenarios (Table 2), resulting from the fitted Weibull degree distribution and the respective BEM. Each cell shows the posterior median and the 95% uncertainty interval of the proportion of recovered MSM. This table corresponds to assuming a maximum number of 350 non-steady partners per year, as a sensitivity analysis to Table A8 that assumed 200 non-steady partners per year and was used for the main text.

| Country | Baseline natural immunity scenario | High natural immunity scenario |
| --- | --- | --- |
| Austria | 1.32% (0.75%, 2.15%) | 1.94% (1.20%, 2.94%) |
| Belgium | 1.58% (0.93%, 2.49%) | 2.91% (1.97%, 4.14%) |
| Bulgaria | 1.32% (0.51%, 2.93%) | 1.76% (0.72%, 3.60%) |
| Croatia | 1.07% (0.42%, 2.22%) | 1.53% (0.66%, 2.92%) |
| Cyprus | 1.12% (0.39%, 2.59%) | 1.53% (0.56%, 3.21%) |
| Czechia | 0.99% (0.44%, 1.90%) | 1.46% (0.72%, 2.58%) |
| Denmark | 1.40% (0.70%, 2.57%) | 1.73% (0.88%, 3.01%) |
| Estonia | 1.10% (0.37%, 2.57%) | 1.76% (0.66%, 3.83%) |
| Finland | 1.44% (0.74%, 2.54%) | 2.10% (1.22%, 3.43%) |
| France | 1.15% (0.82%, 1.56%) | 2.45% (1.90%, 3.14%) |
| Germany | 1.75% (1.43%, 2.12%) | 2.62% (2.23%, 3.06%) |
| Greece | 1.02% (0.48%, 1.91%) | 1.36% (0.71%, 2.29%) |
| Hungary | 1.50% (0.76%, 2.70%) | 1.94% (1.06%, 3.24%) |
| Ireland | 1.22% (0.62%, 2.17%) | 2.34% (1.42%, 3.66%) |
| Italy | 0.81% (0.52%, 1.20%) | 1.67% (1.24%, 2.22%) |
| Latvia | 1.68% (0.71%, 3.80%) | 2.20% (0.97%, 4.55%) |
| Lithuania | 1.09% (0.37%, 2.57%) | 1.78% (0.67%, 3.88%) |
| Luxembourg | 2.14% (0.96%, 4.83%) | 4.07% (2.11%, 7.93%) |
| Malta | 1.22% (0.46%, 2.71%) | 1.64% (0.66%, 3.32%) |
| Netherlands | 1.46% (0.91%, 2.26%) | 2.27% (1.54%, 3.23%) |
| Norway | 1.88% (1.15%, 2.88%) | 3.10% (2.12%, 4.36%) |
| Poland | 1.22% (0.67%, 2.06%) | 1.62% (0.94%, 2.56%) |
| Portugal | 2.01% (1.38%, 2.84%) | 3.18% (2.37%, 4.20%) |
| Romania | 0.77% (0.28%, 1.58%) | 1.34% (0.63%, 2.41%) |
| Slovakia | 1.01% (0.35%, 2.22%) | 1.31% (0.48%, 2.68%) |
| Slovenia | 1.28% (0.50%, 2.80%) | 1.69% (0.69%, 3.39%) |
| Spain | 3.07% (2.49%, 3.75%) | 4.16% (3.50%, 4.92%) |
| Sweden | 1.19% (0.71%, 1.87%) | 1.79% (1.17%, 2.60%) |

**Figure A8.**
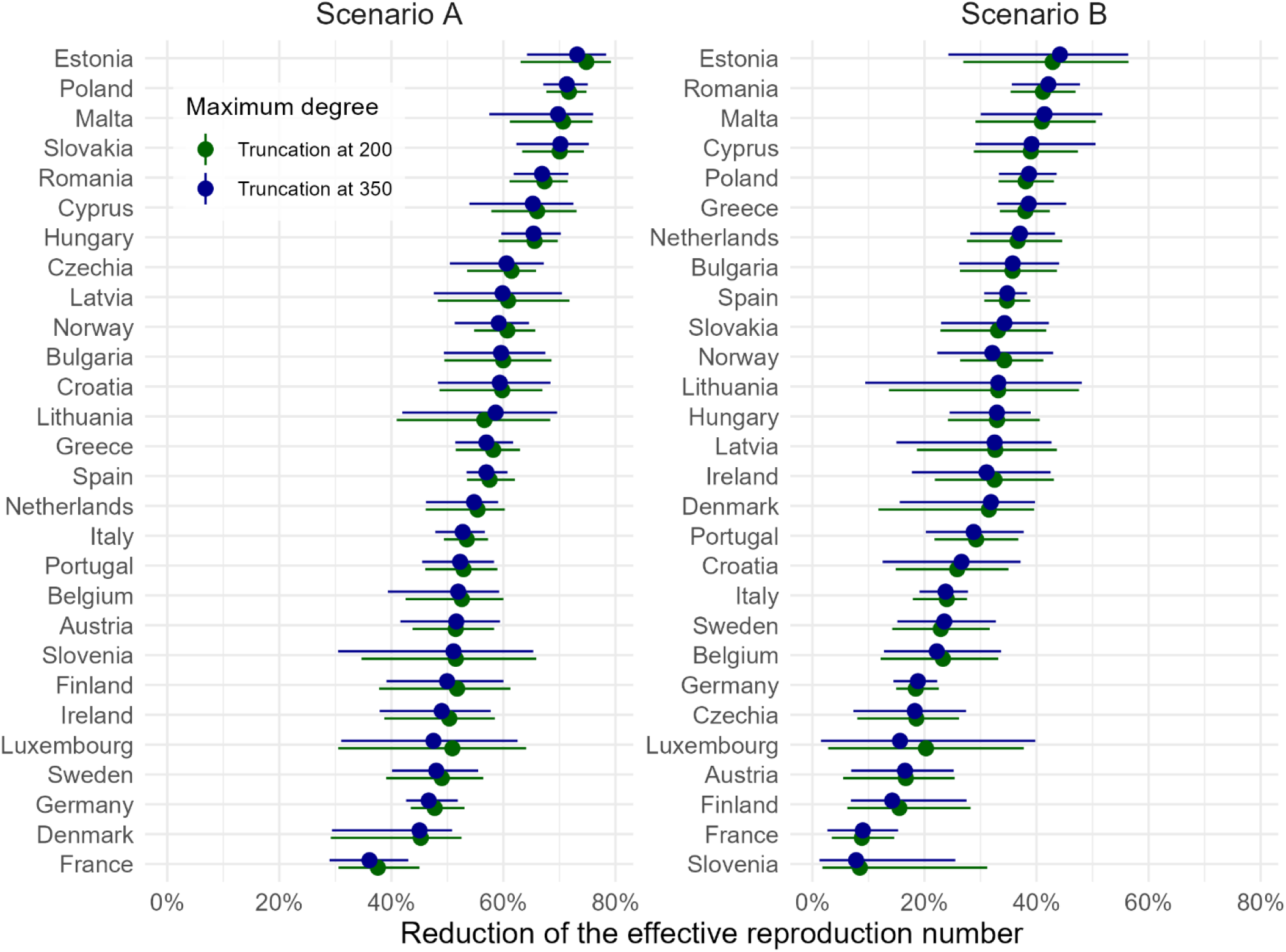
The potential of increasing the mpox vaccine uptake among MSM to reduce transmission, comparing different truncation values for the number of non-steady partners. The reduction of the effective reproduction number ℛ_eff_, obtained by comparing the increased vaccination scenarios A and B with the baseline vaccination scenario. The results are aggregated over the baseline and high natural immunity scenarios as well as aggregated over a range of the effective reproduction number from ℛ_eff_ = 1 to ℛ_eff_ = 4 in the baseline vaccination scenario. The points correspond to the respective median, and the width of the lines shows the 95% uncertainty interval, obtained from 100 posterior samples of the Bayesian mixed-effects logistic regression models of the vaccine and recovery status versus country and number of sexual partners fitted to EMIS-2024 data.

## Appendix C. Incorporating vaccination and recovery into the degree distributions

We model the sexual contact network on each layer as a configuration network model and denote the random variable *D*_all,*l*_ as the number of steady (*l* = 1) and non-steady (*l* = 2) partners of a randomly chosen individual over the last 12 months, which we describe by a discrete Weibull distribution as in [25] and truncated at a maximum degree of *d*_max,1_ = 10 and *d*_max,2_ = 200, respectively. In contrast to *D*_*l*_, which is the degree *among susceptible* individuals on layer *l*, the random variable *D*_all,*l*_ considers contacts *among all individuals* on layer *l*. Non-susceptible (i.e., recovered or having vaccine-induced protection) individuals are effectively removed from the population underlying the epidemic. Hence, the effective reproduction number ℛ_eff_, via the offspring matrix in (1), depends on the degree distribution *D*_*l*_ among susceptible MSM. In the following, we describe how to obtain *D*_*l*_ from *D*_all,*l*_.

As a first step, we focus on layer *l* = 2 and compute Pr[*D*_all,2_ = *x, S*], the probability of an individual to have *x* contacts on layer *l* and to be susceptible. An individual is susceptible if all of the three points below hold true:

1. The individual has no protection against infection from smallpox vaccination. With a probability *p*_spox_ = 14.98%, a randomly chosen MSM is immune against mpox due to historic smallpox vaccination, which is obtained from averaging over the country-specific immunity levels [31] across countries.
2. The individual is not recovered from mpox. For an MSM with *x* non-steady partners over the last 12 months, the BME model fitted to the EMIS-2024 data provides the probability Pr [*R*|*D*_all,2_ = *x*] of being recovered from mpox.
3. The individual has no protection against infection from mpox vaccination. This is the case if the individual is not vaccinated against mpox or if the individual has been vaccinated against mpox but is at risk of a breakthrough infection due to an imperfect vaccine effectiveness. The BME model fitted to the EMIS-2024 data provides the probability Pr[*V*_2_|*D*_all,2_ = *x*] of being vaccinated with two doses, and the conditional probability Pr[*V*_1_|¬*V*_2,base_, *D*_all,2_ = *x*] of being vaccinated with one dose. The probability for one-dose vaccinated is conditional on not having received two doses in the *baseline scenario*, stressed by the “base” subscript for *V*_2,base_. For each of the three scenarios, the unconditional probability of being vaccinated with one dose follows from using the mutual exclusivity (and hence *V*_1_ ∧ ¬*V*_2,base_ = *V*_1_) as

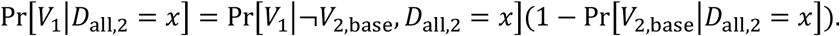

An individual has no vaccine-induced protection against infection if they are unvaccinated, vaccinated once but affected by an imperfect vaccine effectiveness, or vaccinated (with one or two doses) but affected by an imperfect vaccine effectiveness. Hence, the probability of having no vaccine-induced protection follows as

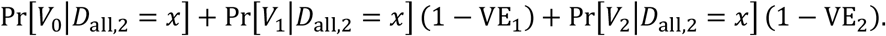

Here, the last two addends are due to the probabilities of breakthrough infections (1 − VE_1_)Pr[*V*_1_|*D*_all,2_ = *x*] and (1 − VE_2_)Pr[*V*_2_|*D*_all,2_ = *x*], where VE_2_ = 82% and VE_1_ = 72% denotes the vaccine effectiveness against infection following two-dose and one-dose vaccination, respectively [22].

By combining the probabilities of the three outcomes above, we obtain that

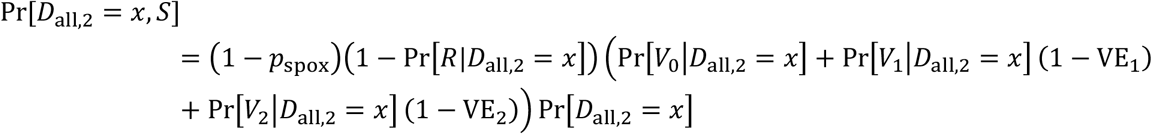

for any degree *x* = 0, …, *d*_max,2_. To reflect that mpox can only spread among susceptible individuals, we adjust the degree distribution *D*_all,2_ of the configuration network model in two steps: First, we remove the half-links starting at any non-susceptible individual, by assigning a zero degree to all individuals that are non-susceptible. We denote the resulting degree distribution by 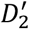, which follows as 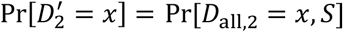 for degrees *x* = 1, …, *d*_max,2_. For *x* = 0, since we remove half-links from those not susceptible, we need to add the probability of an individual not being susceptible

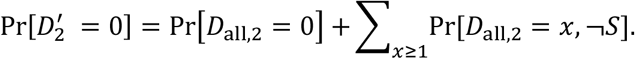

Second, we remove the half-links of susceptible individuals that would be paired with the half-links of non-susceptible individuals (i.e., the half-links reflected by the sum above). The probability *p*_¬*S*,2_ that, on layer 2, a half-link is paired with a half-link of a non-susceptible individual is equal to the fraction of the number of half-links to non-susceptible individuals divided by the number of all half-links. Hence, it follows that

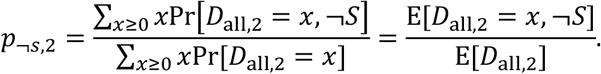

For *x* ≥ 1, it holds that

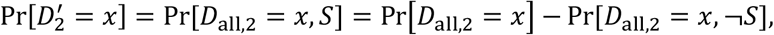

which implies that

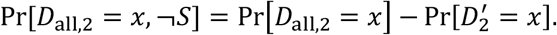

Thus, we obtain for the expected values (where the expectation is taken with respect to the random variable *D*_all,2_) that

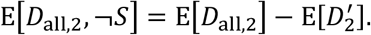

This yields the probability

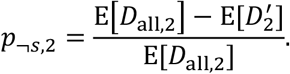

Finally, with the probability *p*_*S*,2_ = 1 − *p*_¬*s*,2_ of a half-link being connected to the half-link of a susceptible individual on layer 2, we obtain the degree distribution *D*_2_ among susceptible individuals as a mixture of binomial distributions

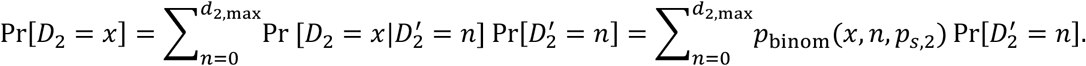

Here, *p*_binom_(*x, n, p*_*s*,2_) denotes the probability of *x* successes of a binomial distribution with *n* trials and success probability *p*_*s*,2_, i.e.,

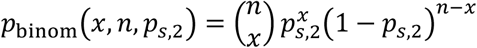

for *x* ≤ *n*, and we define *p*_binom_(*x, n, p*_*s*,2_) = 0 if *x* > *n*, since the number of successes cannot be larger than the number of trials.

For the steady layer *l* = 1, provided the two network layers are independent, we obtain the distribution of the number of steady partners of a susceptible individual as

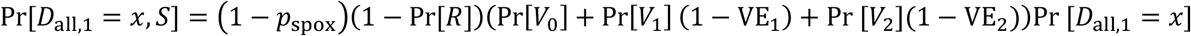

for *x* = 0, …, *d*_max,1_, Here, Pr[*V*_2_] follows from the law of total probability as

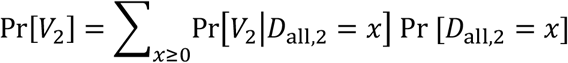

and the probabilities Pr[*V*_1_], Pr[*V*_0_], Pr[*R*] are obtained analogously. We obtain the degree distribution *D*_1_ from Pr[*D*_all,1_ = *x, S*] in the same way as we obtained *D*_2_ from Pr[*D*_all,2_ = *x, S*], outlined above.

For a few countries and few posterior samples, the probability of being vaccinated in the increased vaccination scenario (A or B) is smaller than in the baseline scenario in the tail of the non-steady partnership distribution. In this case, we set the respective vaccination probability of the increased vaccination scenario to the vaccination probability in the baseline scenario.

## Appendix D. Reproduction number estimates

Figure A9 shows the time-varying reproduction number for France, Germany, and Spain between 1 August 2024 – 1 August 2026, estimated with the EpiEstim R package by Cori et al. [32, 33] based on the data available at Joint ECDC-WHO Regional Office for Europe Mpox Surveillance Bulletin [3]. We focus on these three countries since they have the most mpox cases in the EU/EEA in the considered time period and are less affected by data sparsity. In line with Miura et al. [34], we set the mean and standard deviation of the serial interval to 10.1 days and 3.5 days, respectively.

**Figure A9.**
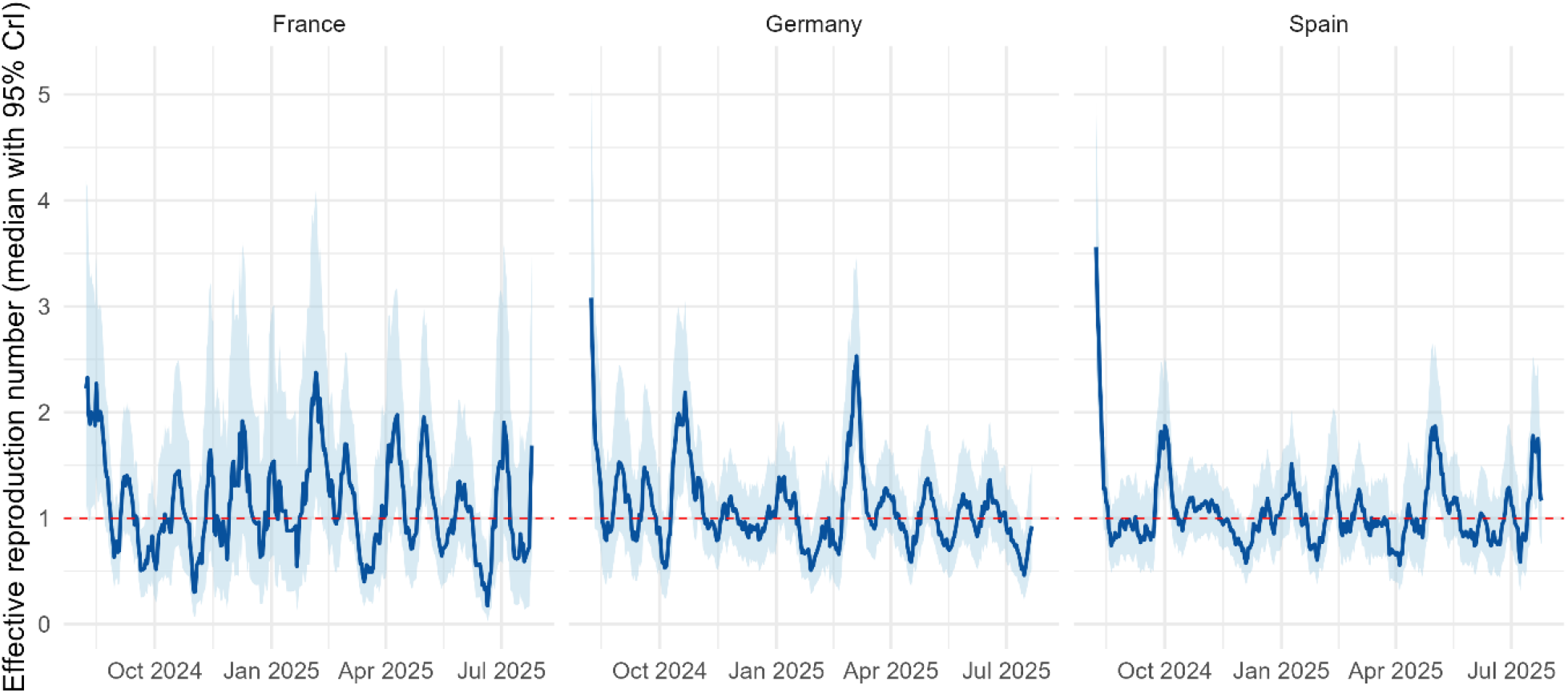
The effective reproduction number from August 2024 – August 2025 in the three EU/EEA countries with the most mpox cases. The blue line shows the median posterior estimate, and the shaded blue area corresponds to the 95% credible interval. The dashed red line indicates an effective reproduction number ℛ_eff_ of 1.

Figure A9 motivates our choice to fit the reproduction number in the baseline scenario to a range of values from ℛ_eff_ = 1 to ℛ_eff_ = 4. Across all countries, scenarios and values for ℛ_eff_, the resulting median value for the infection rate multiplier, fitted via bisection, was *α* = 0.86 (95% UI: 0.15-2.21).

Figure A10 shows the expected reduction of the reproduction number disaggregated by the effective reproduction number ℛ_eff_ in the baseline scenario. (Figure 4 in the main text shows these results aggregated across ℛ_eff_ = 1 to ℛ_eff_ = 4.) Figure A10 shows that the effective reproduction number ℛ_eff_ in the baseline scenario does not have a substantial impact on the relative reduction of the reproduction number,

**Figure A10.**
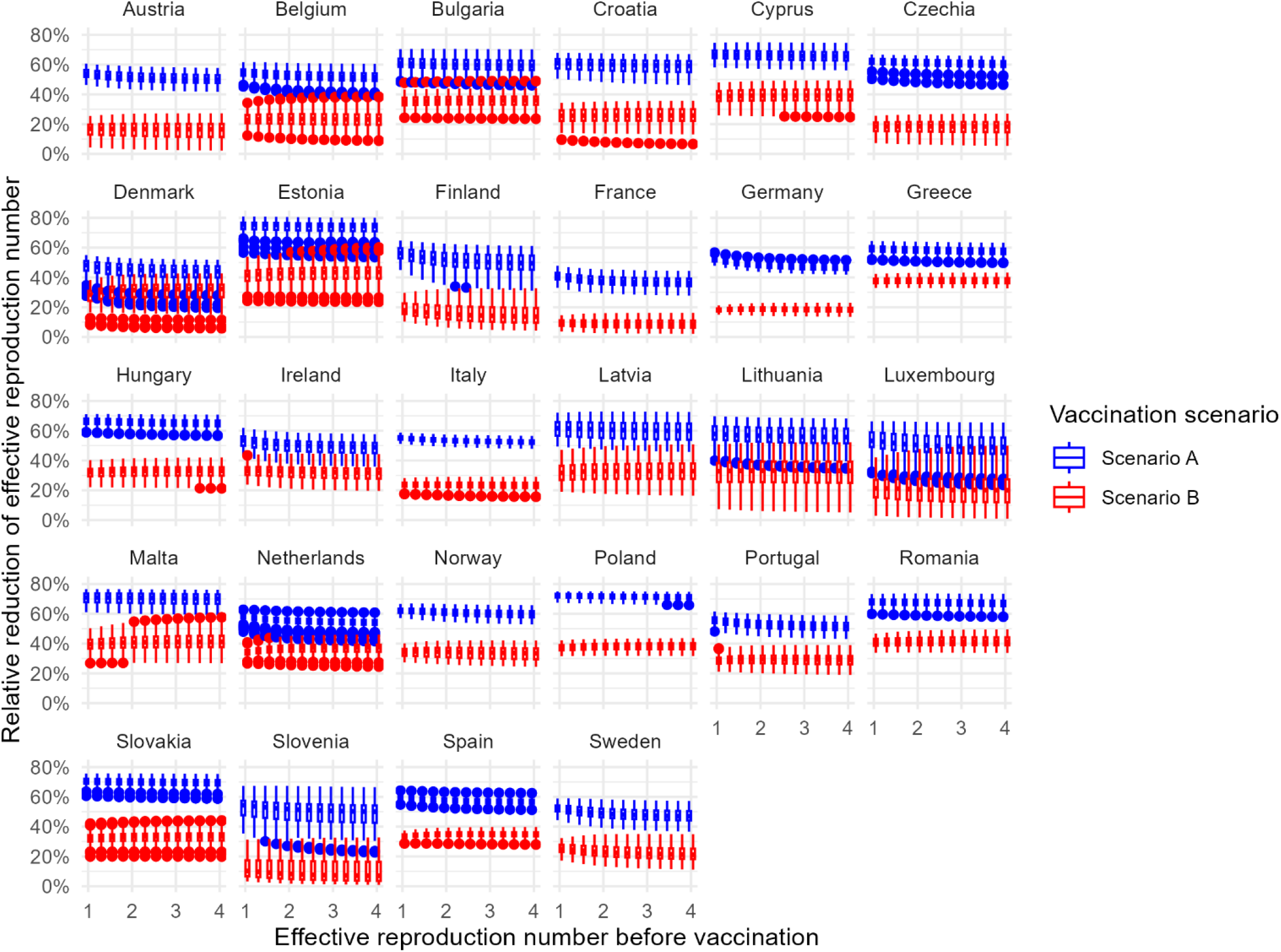
The potential of increasing the mpox vaccine uptake among MSM to reduce transmission versus the effective reproduction number in the baseline scenario. The reduction of the effective reproduction number ℛ_eff_, obtained by comparing the increased vaccination scenarios A and B with the baseline vaccination scenario. The results are aggregated over the baseline and high natural immunity scenarios. The boxplots are obtained from summarising the results from 100 posterior samples of the Bayesian mixed-effects logistic regression models of the vaccine and recovery status versus country and number of sexual partners fitted to EMIS-2024 data.

## Appendix E. Scenario and sensitivity analyses of the vaccination impact on the effective reproduction number

**Figure A11.**
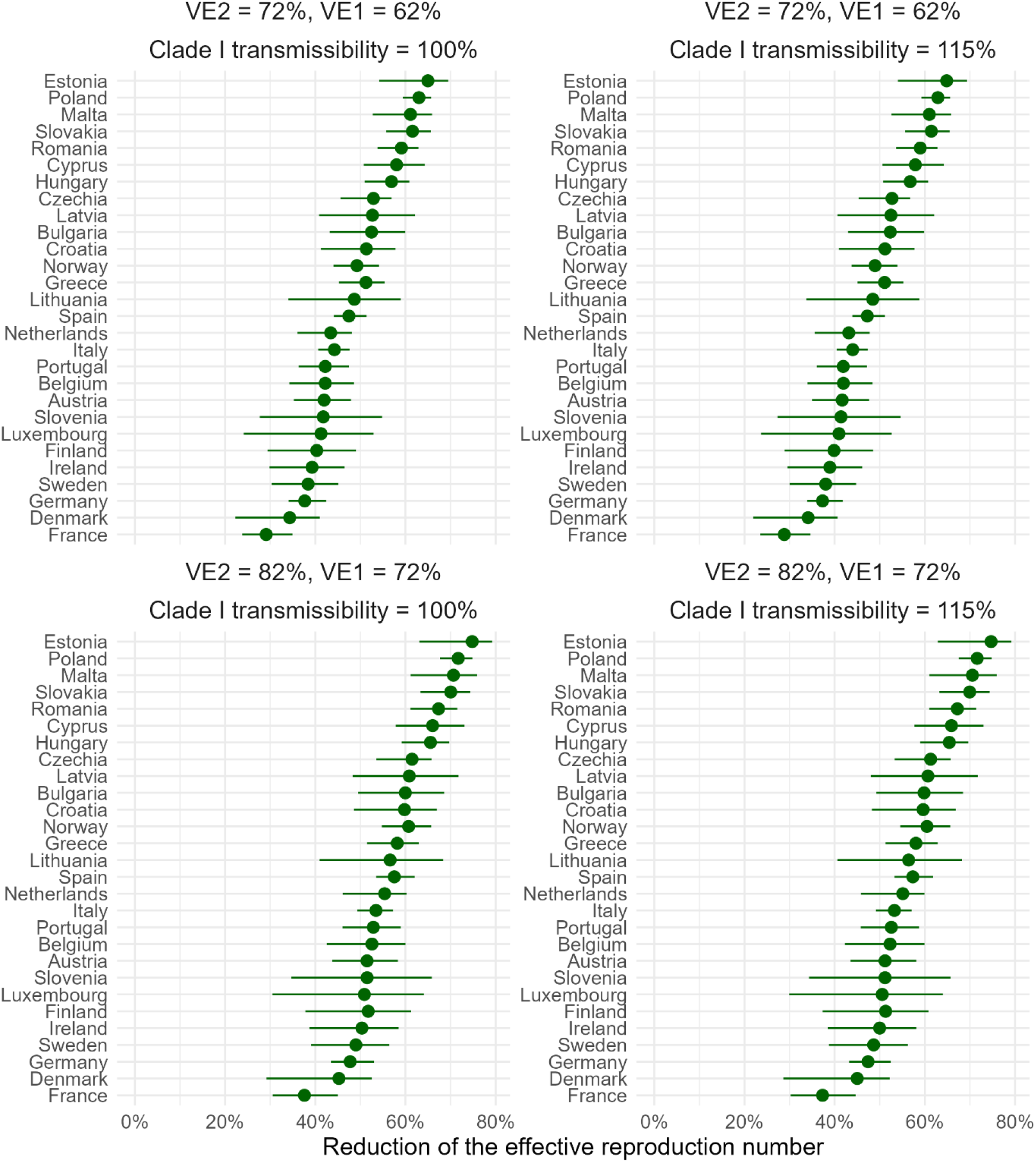
The potential of increasing the mpox vaccine uptake among MSM (scenario A) to reduce transmission versus the MPXV clade I transmissibility and the vaccine effectiveness VE_2_ and VE_1_ following two doses and one dose vaccination. The reduction of the effective reproduction number ℛ_eff_, obtained by comparing the increased vaccination scenario A with the baseline vaccination scenario. The transmissibility of MPXV clade I is given relative to the transmissibility of clade II, hence, 100% corresponds to an equivalent transmissibility of the two clades. The results are aggregated over the baseline and high natural immunity scenarios as well as aggregated over a range of the effective reproduction number from ℛ_eff_ = 1 to ℛ_eff_ = 4 in the baseline vaccination scenario. The points correspond to the respective median, and the width of the lines shows the 95% uncertainty interval, obtained from 100 posterior samples of the Bayesian mixed-effects logistic regression models of the vaccine and recovery status versus country and number of sexual partners.

**Figure A12.**
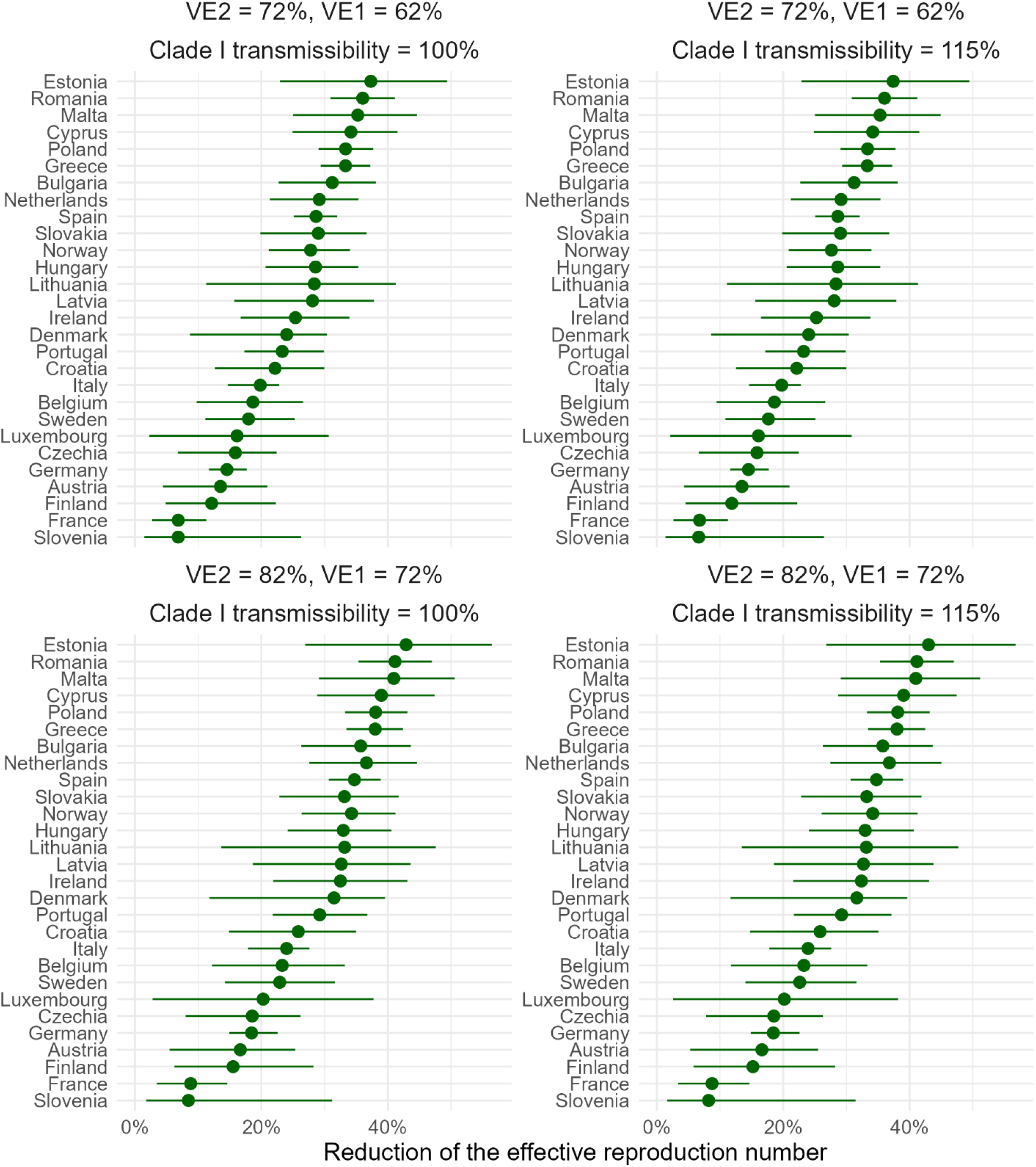
The potential of increasing the mpox vaccine uptake among MSM (scenario B) to reduce transmission versus the MPXV clade I transmissibility and the vaccine effectiveness VE2 and VE1 following two doses and one dose vaccination. The reduction of the effective reproduction number ℛ_eff_, obtained by comparing the increased vaccination scenario B with the baseline vaccination scenario. The transmissibility of MPXV clade I is given relative to the transmissibility of clade II, hence, 100% corresponds to an equivalent transmissibility of the two clades. The results are aggregated over the baseline and high natural immunity scenarios as well as aggregated over a range of the effective reproduction number from ℛ_eff_ = 1 to ℛ_eff_ = 4 in the baseline vaccination scenario. The points correspond to the respective median, and the width of the lines shows the 95% uncertainty interval, obtained from 100 posterior samples of the Bayesian mixed-effects logistic regression models of the vaccine and recovery status versus country and number of sexual partners.

**Figure A13.**
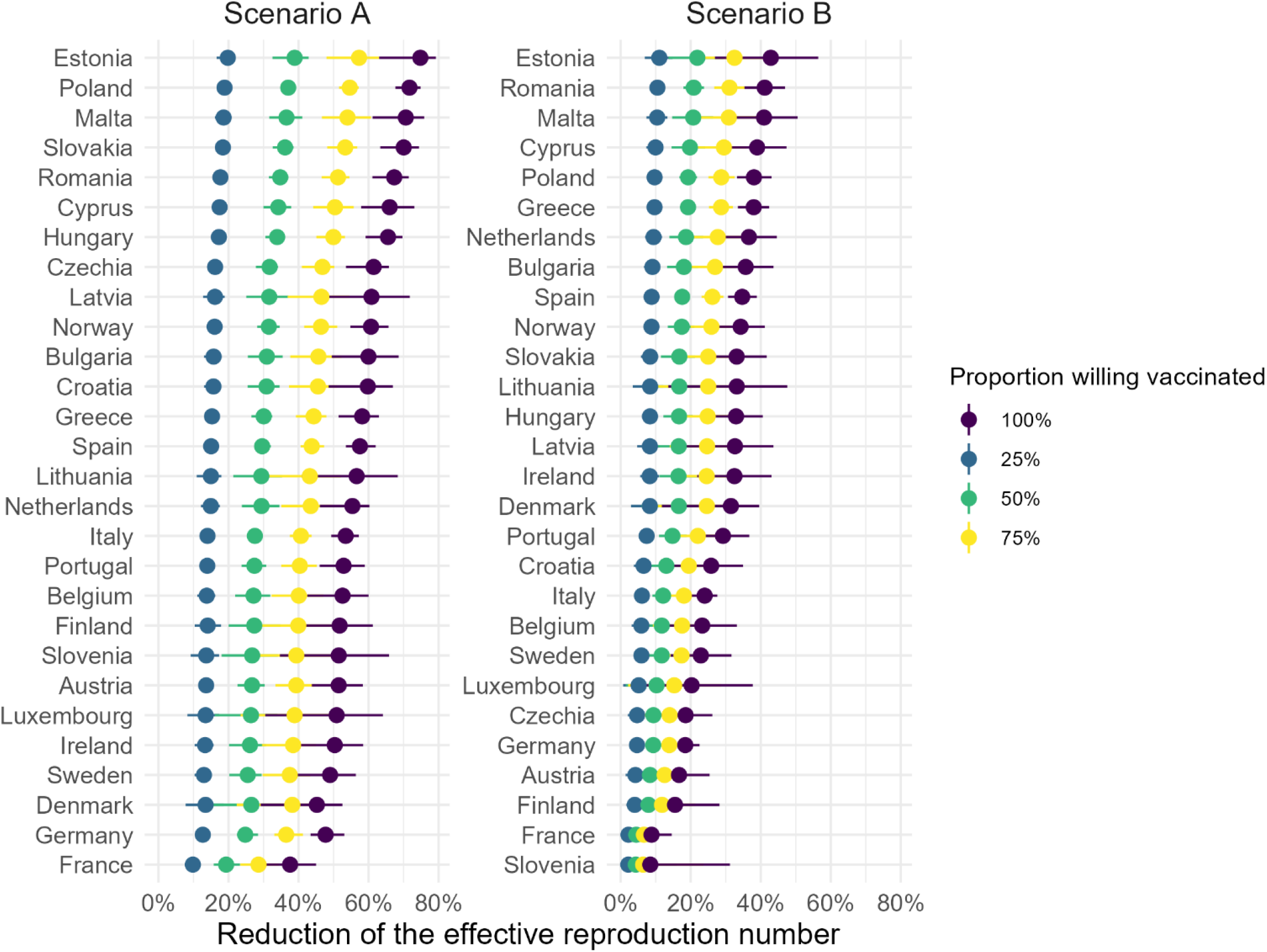
The potential of increasing the mpox vaccine uptake among MSM to reduce transmission. The reduction of the effective reproduction number ℛ_eff_, obtained by comparing the increased vaccination scenarios A and B with the baseline vaccination scenario. Here, the ‘proportion willing vaccinated’ equals the individuals that are additionally vaccinated divided by the number of individuals that would like to get vaccinated in the respective scenario. (Hence, proportion willing vaccinated of 100% corresponds to Scenario A and B as in Figure 4.) The results are aggregated over the baseline and high natural immunity scenarios as well as aggregated over a range of the effective reproduction number from ℛ_eff_ = 1 to ℛ_eff_ = 4 in the baseline vaccination scenario. The points correspond to the respective median, and the width of the lines shows the 95% uncertainty interval, obtained from 100 posterior samples of the Bayesian mixed-effects logistic regression models of the vaccine and recovery status versus country and number of sexual partners.

## Footnotes

1 While defining a “typical” individual is straightforward for homogeneous populations, one must take more caution for heterogenous populations – such as MSM sexual networks.

2 More precisely, one could write 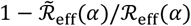, since the reproduction numbers depend on the infection rate multiplier *α*. However, for ease of notation, we drop the argument *α*. The variation with respect to *α* is incorporated in the uncertainty bars of the results.

